# Population Normalization in SARS-CoV-2 Wastewater-Based Epidemiology: Implications from Statewide Wastewater Monitoring in Missouri

**DOI:** 10.1101/2022.09.08.22279459

**Authors:** Chenhui Li, Mohamed Bayati, Shu-Yu Hsu, Hsin-Yeh Hsieh, Wilfing Lindsi, Anthony Belenchia, Sally A. Zemmer, Jessica Klutts, Mary Samuelson, Melissa Reynolds, Elizabeth Semkiw, Hwei-Yiing Johnson, Trevor Foley, Chris G. Wieberg, Jeff Wenzel, Terri D. Lyddon, Mary LePique, Clayton Rushford, Braxton Salcedo, Kara Young, Madalyn Graham, Reinier Suarez, Anarose Ford, Dagmara S. Antkiewicz, Kayley H. Janssen, Martin M. Shafer, Marc C. Johnson, Chung-Ho Lin

## Abstract

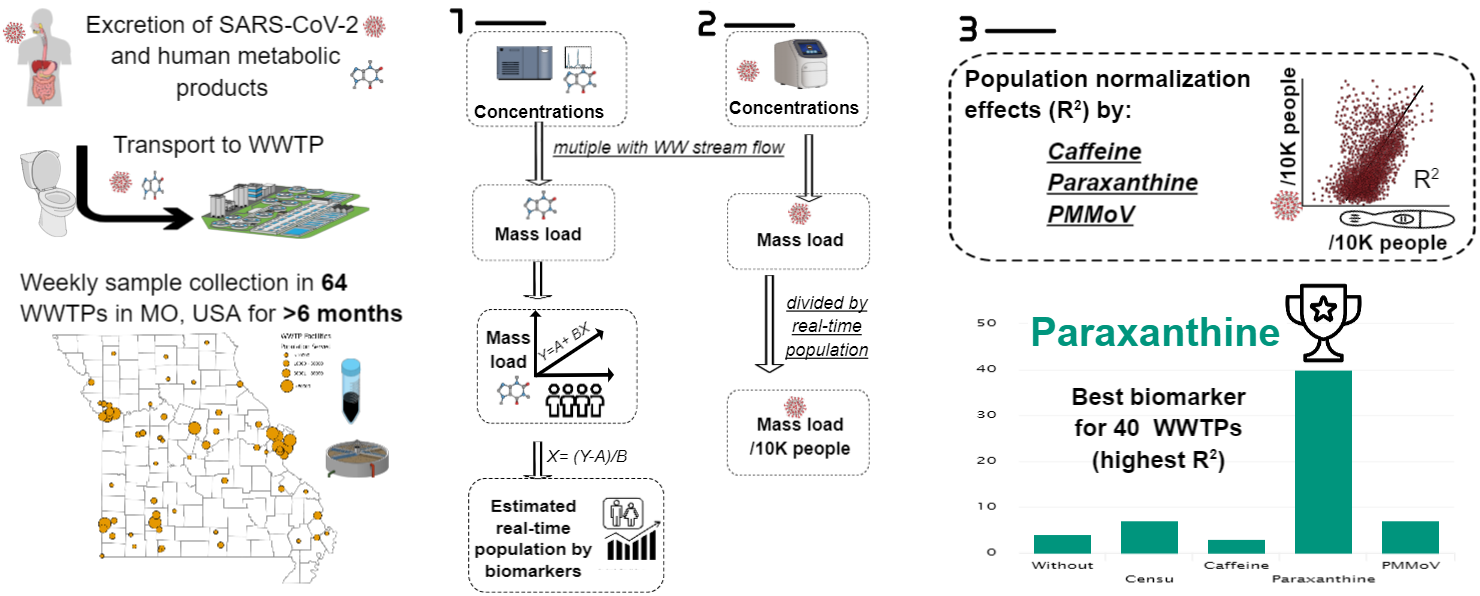

The primary objective of this study was to identify a universal wastewater biomarker for population normalization for SARS-CoV-2 wastewater-based epidemiology (WBE). A total of 2,624 wastewater samples (41 weeks) were collected weekly during May 2021-April 2022 from 64 wastewater facilities across Missouri, U.S. Three wastewater biomarkers, caffeine and its metabolite, paraxanthine, and pepper mild mottle virus (PMMoV), were compared for the population normalization effectiveness for wastewater SARS-CoV-2 surveillance. Paraxanthine had the lowest temporal variation and strongest relationship between population compared to caffeine and PMMoV. This result was confirmed by data from ten different Wisconsin’s WWTPs with gradients in population sizes, indicating paraxanthine is a promising biomarker of the real-time population across a large geographical region. The estimated real-time population was directly compared against the population patterns with human movement mobility data. Of the three biomarkers, population normalization by paraxanthine significantly strengthened the relationship between wastewater SARS-CoV-2 viral load and COVID-19 incidence rate the most (40 out of 61 sewersheds). Caffeine could be a promising population biomarker for regions where no significant exogenous caffeine sources (e.g., discharges from food industries) exist. In contrast, PMMoV showed the highest variability over time, and therefore reduced the strength of the relationship between sewage SARS-CoV-2 viral load and the COVID-19 incidence rate, as compared to wastewater data without population normalization and the population normalized by either recent Census population or the population estimated based on the number of residential connections and average household size for that municipality from the Census. Overall, the findings of this long-term surveillance study concluded that the paraxanthine has the best performance as a biomarker for population normalization for SARS-CoV-2 wastewater-based epidemiology.

## 1. INTRODUCTION

Wastewater-based epidemiology (WBE) provides human activity information within sewershed boundaries by relating concentrations of chemical and biological “waste” materials in wastewater influent to population-scale use, consumption of pharmaceuticals, illicit drugs, or rates of exposure to industrial chemicals.^1^ Over the last few years, WBE applications have been emerging in infectious diseases or pathogens and antibiotic resistance, especially since the COVID-19 pandemic. The WBE is widely recognized as a valuable tool for monitoring community trends in COVID-19 with the advantages of providing an efficient and representative population-pooled sample, and complementing community data, especially where timely COVID-19 clinical testing is underused or unavailable.^2^ Moreover, WBE reveals the underestimation of COVID-19 clinical testing because SARS-CoV-2 is shed by people with and without symptoms.^3,4^ The WBE can also provide a useful early warning of the emergence or re-emergence of COVID-19 in a community, and afford timely insights for public health interventions, with previous studies showing that SARS-CoV-2 could be detected in wastewater up to two weeks before the cases were reported.^5^ Furthermore, wastewater surveillance can be implemented in most communities since municipal wastewater collection systems serve nearly 80 percent of U.S. households.^6^

The utility of WBE for cost-effective surveillance of SARS-CoV-2 levels in communities was recognized early in the COVID-19 pandemic. However, substantive uncertainty remains in how best to account for the contributing population and fecal strength. Robust population biomarkers are necessary to determine the SARS-CoV-2 load per capita, such that 1) the changes in wastewater virus concentration due to the dilution (e.g., increased volume resulting from major rainfall events or receiving additional discharge from industrial or natural resources) and 2) population dynamics, are accounted for. Wastewater viral loads change with the variations in daily wastewater flow, the proportion of industrial discharge, and the gross proportions of solids, which can be influenced by the design and condition of the wastewater collection system.^7^ For example, SARS-CoV-2 concentration is influenced if the WWTP is receiving wastewater from a combined sewer system that collects domestic wastewater and rainwater runoff in the same pipe; the weather effects, such as precipitation and infiltration/inflow into the sewers, impact the human fecal concentration. In addition, the population size contributing to the sewershed is expected to change over the surveillance period (due to deaths, births, tourism, weekday commuters, pandemic lockdown, temporary workers, etc.).^1^ Therefore, temporal variation in wastewater volume/strength and population size must be better accounted for to make results more comparable over time.^8^ However, population normalization approaches are still under development, and few studies have compared approaches/biomarkers systematically for optimization of wastewater SARS-CoV-2 data for predicting the clinical prevalence of COVID-19.^9^

The population variation is usually monitored and normalized using human fecal or urine biomarkers. Suitable human population normalization controls should meet certain criteria.^8,10,11^ For example, the chemical biomarkers should be specific to human metabolism, excreted into sewage, exogenous sources are minimal, minimal intra- and inter-individual variance in daily excretion, and levels in raw sewage well above the method detection limit. Furthermore, the biomarkers must be stable in the wastewater for a reasonable long time (e.g., during the transport from the toilet to the sampling point and during sampling, storage, and analysis).^12^ In addition, there should be low variance in the per capita daily excretion and not be affected by environmental variables such as season, weather, or geographic location.^8^

Some commonly measured wastewater properties/chemicals, such as chemical and biological oxygen demand, and total nitrogen and phosphorus, have been explored for wastewater population indicators.^13,14^ The disadvantage of these environmental parameters is that they are highly influenced by wastewater composition (i.e., industrial, domestic, or mixed) since they are not only shed by humans but also from exogenous sources such as food waste processed by garbage disposals and fertilizer runoff.^8^ Ammonium originates from the breakdown of urea ^15^ and is introduced via toilets and routinely measured by WWTP as a water quality parameter, which is supposed to be less affected by non-human sources than chemical or biological oxygen demand and total phosphorous.^16^ However, Sweetapple et al. found that population normalization by orthophosphate and ammonium did not result in improvement of correlations between wastewater SARS-CoV-2 data and indicators of COVID-19 prevalence.^14^

In addition, a variety of endogenous and exogenous human biomarkers that can be measured directly in wastewater samples have been evaluated to estimate their human fecal content. These markers include but are not limited to: (1) bacteria/viruses or molecules that are ubiquitous in human intestinal tracts, such as cross-assembly phage, human ribonuclease P,^17^ and *Bacteroides* HF183;^18^ (2) an exogenous substance (or its’ metabolite) after intentional consumption of a substance (i.e., personal care products, food additives and dietary supplements such as carbamazepine and gabapentin,^19^ artificial sweeteners,^20^ and caffeine and its metabolite (paraxanthine),^10^ pepper mild mottle virus (PMMoV);^18^ and (3) endogenous compounds that are produced naturally in the body such as creatinine, cholesterol and its metabolite coprostanol, cortisol, and serotonin metabolite 5-hydroxyindoleacetic acid (5-HIAA).^11^

Pepper mild mottle virus (PMMoV), a virus ingested with pepper-containing food, has been widely measured and frequently used for normalizing SARS-CoV-2 concentration data. The PMMoV is one of the widely used normalization biomarkers in wastewater SARS-CoV-2 surveillance because it is believed to be present in high concentrations in wastewater, introduced into the human body through the diet, and has the potential to serve as an RNA recovery control since it is a single-stranded RNA virus.^21,22^ D’Aoust et al. found PMMoV to be superior to HF183 *Bacteroides* 16S ribosomal rRNA and eukaryotic 18S rRNA, as PMMoV showed more reproducibility within and between WWTPs^18^. Creatinine, the endogenous nitrogenous waste product, has been used to normalize the concentrations of other urinary excretion products to account for urine dilution in clinical chemistry and is recommended as a possible biomarker for estimating the population.^23^ The serotonin metabolite, 5-HIAA, has also been evaluated as a wastewater marker, and was reported to be more stable within sewer systems than cortisol and androstenedione.^24^

Caffeine (1,3,7-trimethylxanthine) is one of the world’s most widely consumed dietary ingredients, found in many globally popular products, including tea, cola and energy drinks, and in some medications and nutritional supplements. Still, the most important source of this alkaloid is coffee.^25^ The excretion of the metabolite of caffeine, paraxanthine (1,7-dimethylxanthine), was less affected by the genetic-based variation in pharmacokinetics than the parent compound was, therefore, suggested as a potential biomarker for dietary caffeine intake.^26^ Chemicals (e.g., paraxanthine) involved in endogenous metabolism (products of biosynthesis or catabolism) avoid xenobiotics’ problems for use as proxy measures for population since their association with per capita activities has higher fidelity.^23^ Caffeine and paraxanthine are easy to detect due to the high concentration levels (μg/L) in untreated wastewater,^27^ and are stable in wastewater samples stored at 4°C,^28^ which makes them ideal biomarkers. However, more research was needed for caffeine and paraxanthine application in the concept of WBE.

In a previous study, we compared five biomarkers, PMMoV, creatinine, 5-HIAA, caffeine, and paraxanthine, based on two weeks’ data, for their utility in normalizing SARS-CoV-2 loads and found caffeine and especially paraxanthine were the most reliable population biomarkers.^29^ This study extended the investigation to the long-term weekly monitoring of 64 wastewater treatment plants (WWTPs) across Missouri for more than six months to compare the utility of caffeine, paraxanthine, and PMMoV as population biomarkers across seasons and distinct geographical areas with contrasting sewershed sizes.

## 2. MATERIALS AND METHODS

### 2.1 Wastewater sampling and clinical COVID-19 case

Triplicates of 50 mL of 24-hr composite influent (before primary treatment) samples were collected once per week from 64 WWTPs in Missouri (Fig. 1), and a total of 41 weeks from May 2021 to April 2022 (weeks of 05/10/2021, 05/24/2021, 06/28/2021, and consecutively from weeks of 07/19/2021 to 04/10/2202). A majority of samples were collected as 24-hr time-proportional composites and only 6 WWTPs were collected as 24-hr flow-proportional composites. Each WWTP collected samples on the same day of the week during the study period. The first half of the sampling period was dominated by the Delta variant, and the second half was dominated by the Omicron variant. In total, 2624 wastewater samples were collected.

**Fig. 1.**
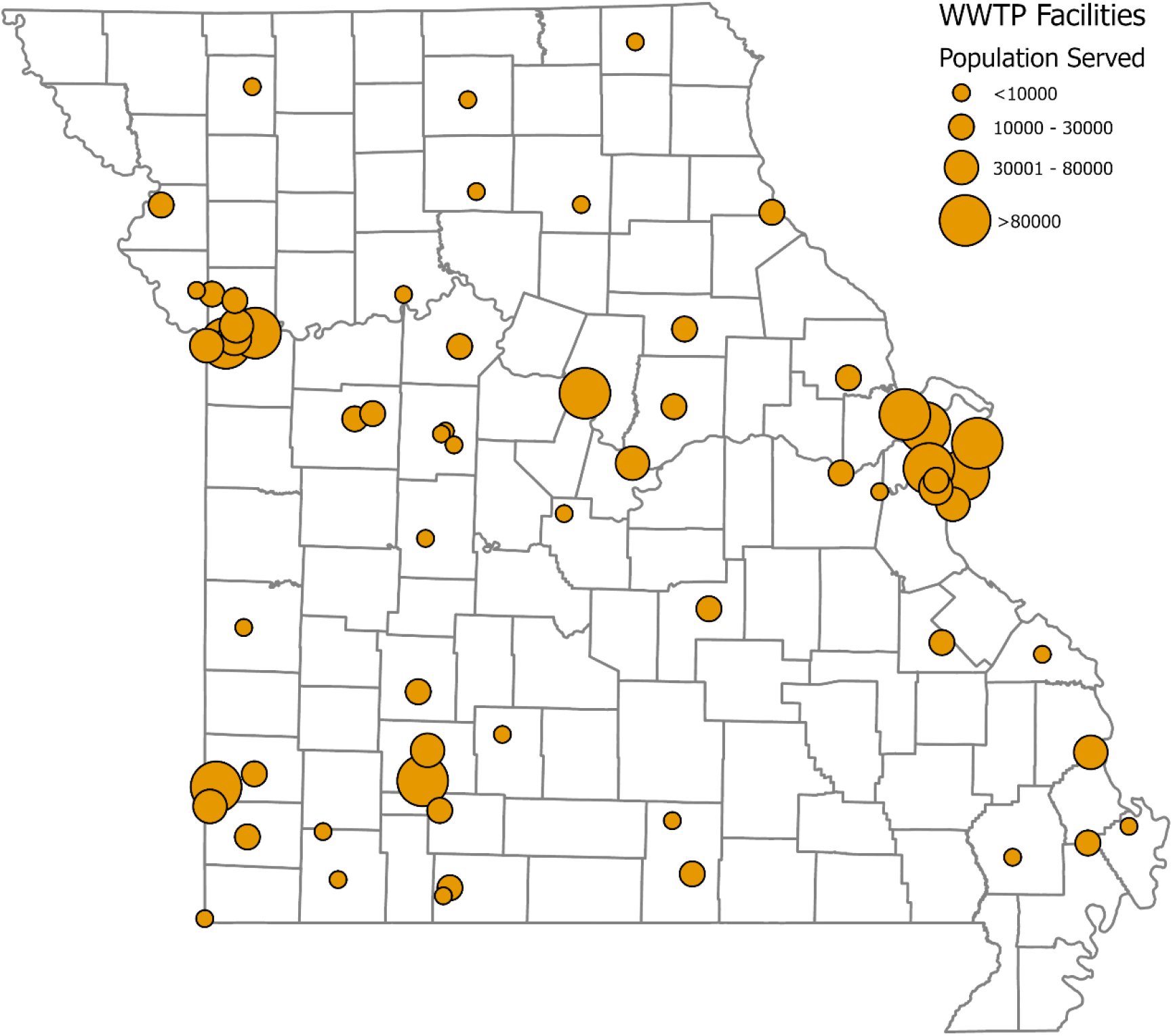
A total of 2624 wastewater samples were collected from 64 wastewater treatment plants (WWTPs) across Missouri, USA.

Wastewater samples were transported in insulated shippers with ice packs to the University of Missouri within 24 hr from the collection and then stored at 4°C until extraction. WWTPs reported their 24-hr flow rates and weekly new COVID-19 cases were provided by the Missouri Department of Health and Senior Services (DHSS). Weekly clinical COVID-19 cases for each WWTP service area were obtained by matching clinically confirmed COVID-19 case data (georeferenced using home address) to WWTPs sewershed boundaries in ArcGIS. Sewershed boundaries were either provided by the municipality or, in many cases (12 WWTPs), the municipality did not have geospatial data delineating their sewershed boundaries, so municipal boundaries were used to approximate a service area.

The 64 WWTPs cover urban, semirural, and rural locations throughout Missouri with the sewershed population ranging from 900 to 490,000. Sewershed size (metadata population) was either provided by the WWTP or estimated based on the number of residential connections reported in a WWTP’s discharge permit and the average household size for that municipality from the most recent U.S. Census at the time the facility began sampling. Based on the population served by the 64 WWTPs, there is the potential to monitor 50% of the 6.15 million Missouri population.^30^ A summary of each facility, including the population served and the locations, is provided in Table S1. Additionally, ten wastewater composite samples collected from WWTPs in Wisconsin during the week of 06/07/2021 were utilized as a data set for model evaluation and validation.

### 2.2 Mobility data

Daily data on mobility (driving and walking) were downloaded for Columbia, MO, between 01/13/2020 and 04/13/2022 from Apple Mobility Trends Reports^31^. Apple mobility data has no demographic information about the users. The original mobility indices were relative percentages to the reference date on 01/13/20, which were scaled to the maximum observed during the study period.

### 2.3. Wastewater concentrations of caffeine and paraxanthine

#### 2.3.1. Extraction of caffeine and paraxanthine

For all 2,624 wastewater samples, 1.5 ml was centrifuged at 10000 rpm for 10 minutes, and 0.75 ml supernatant was extracted and mixed with 0.75 ml ammonium acetate buffer (10 mM ammonium acetate and 0.1% formic acid in water). Then, 10 µL of formic acid was added to precipitate the large molecules in the wastewater, followed by spiking Caffeine-C^13^ to evaluate the recovery of endogenous caffeine. For improved sample storage stability, the 0.75 ml ammonium acetate buffer was replaced by 0.75 ml 100% methanol starting from December 2021. The two preparation methods (ammonium acetate buffer and methanol) were compared, and no significant difference in caffeine and paraxanthine concentrations was found before the transition (Table S2). Finally, the mixture was filtered through 0.2 µm PTFE filters (13mm) (Waters, USA) before the liquid chromatography with tandem mass spectrometry (LC-MS/MS) analysis.

#### 2.3.2. Liquid chromatography-tendon mass spectrometry analysis

The quantification of caffeine and paraxanthine was performed on a Waters Alliance 2695 High Performance Liquid Chromatography (HPLC) system coupled with Waters Acquity TQ triple quadrupole mass spectrometer (MS/MS). The analytes were separated on a Phenomenex (Torrance, CA) Kinetex C18 (100mm x 4.6 mm; 2.6 µm particle size) reverse-phase column. The mobile phase consisted of 10 mM ammonium acetate and 0.1% formic acid in water (A) and 100% acetonitrile (B). The gradient conditions were 0 – 0.3 min, 2% B; 0.3-7.27 min, 2-80% B; 7.27-7.37 min, 80-98% B; 7.37-9.0 min, 98% B; 9-10 min 98-2% B; 10.0 – 15.0 min, 2% B at a flow rate of 0.5 mL/min. The ion source in the MS/MS system was electrospray ionization (E.I.) operated in positive ion mode with a capillary voltage of 1.5 kV. The temperature of the ionization source was 150°C, and that of the desolvation zone, 450°C. The optimized collision energy, cone voltage, and molecular and product ions of the biomarkers are summarized in Table 1.

**Table 1.** Summary of the optimized LC-MSMS Parameters for chemical population biomarkers.

| No. | Compound | RT | ES | MS1 | MS2 | Cone Voltage | Collision Energy |
| --- | --- | --- | --- | --- | --- | --- | --- |
| 1 | Caffeine | 6.273 | ES+ | 195.05 | 138.12 | 45 | 22 |
| 2 | Caffeine-<sup>13</sup>C<sub>3</sub> | 6.167 | ES+ | 198.04 | 140.07 | 45 | 22 |
| 3 | Paraxanthine | 5.715 | ES+ | 181.06 | 124.11 | 45 | 22 |

### 2.4.1 Wastewater concentrations of SARS-CoV-2 and PMMoV

#### 2.4.1. RNA extraction

Samples from the weeks of 09/13/2021 to 04/10/2022 were processed for both SARS-CoV-2 and PMMoV quantification. First, 50-ml raw wastewater samples were spun at 2000 ×g for 5 min to remove large particulates, then vacuum filtered through a 0.22 μm filter (Millipore cat# SCGPOO525). Then, 35.5 mL of filtered wastewater was mixed with 12 mL of 50% (W/V) polyethylene glycol (PEG, Research Products International, cat# P48080) and 1.2M NaCl, followed by equilibration for 2 h at 4°C, all done on the day of sample receipt. Afterwards, samples were centrifuged at 12,000 x g for 2 h. RNA was extracted from the pellet using the Qiagen Viral RNA extraction kit following the manufacturer’s instructions after removing the supernatant. RNA was eluted in a final volume of 56 μL and stored at −80°C if it was not processed immediately.

#### 2.4.2. RT-qPCR

The extracted RNA was used to perform RT-qPCR quantification of the genetic material of SARS-CoV-2 and PMMoV, separately. SARS-CoV-2 was quantified using the primer and control sets described in Robinson et al.,^32^ and PMMoV was quantified using the primer sets described in Hsu et al.^29^ A plasmid carrying a PMMoV gene along with an N gene fragment was constructed, purified from *Escherichia coli*, and used as standards for the RT-qPCR assay. Final RT-qPCR one-step mixtures consisted of 5 µL TaqPath 1-step RT-qPCR Master Mix (Thermo Fisher), 500 nM of each primer, 125 nM of each TaqMan probes, 5 µl of wastewater RNA extract, and RNase/DNase-free water to reach a final volume of 20 µL. All RT-qPCR assays were run in duplicate on a 7500 Fast real-time qPCR instrument (Applied Biosystems). The reactions were initiated with one cycle of UNG incubation at 25°C for 2 min and then one cycle of reverse transcription at 50°C for 15 min, followed by one cycle of activation of DNA polymerase at 95°C for 2 min and then 45 cycles of 95°C for 3 sec for DNA denaturation and 55°C for 30 sec for annealing and extension. The data would be collected at the step of 55°C extensions. The concentration of SARS-CoV-2 and PMMoV genomes in each sample, [*N1.N2*]_*SARS*_ or [PMMoV] (copies/L), was calculated using Eq. 1.

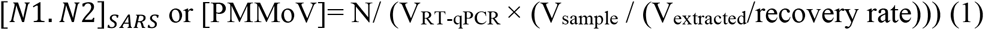

where N (copies/reaction) is the gene copies detected in each RT-qPCR reaction, V_RT-qPCR_ is the volume of RNA used for RT-qPCR (5 µL), V_sample_ is the wastewater sample volume initially used for the concentration step (35.5 mL), and V_extracted_ is the total volume of nucleic acid extracted (56 µL).

### 2.5 Statistical analyses

The statistical analyses included the following three steps: 1) estimation of the real-time population using the wastewater biomarker mass loads; 2) population normalization for wastewater SARS-CoV-2 viral load and COVID-19 case to wastewater SARS-CoV-2 viral concentration (copies/week/10,000 people) and COVID-19 incidence rate (case/week/10,000 people), and 3) comparison of the strength of the relationships between population normalized wastewater SARS-CoV-2 viral load and COVID-19 incidence rate under different normalization scenarios.

#### 2.5.1. Real-time population estimation using wastewater biomarker loads

All the data analyses were conducted using the R program.^33^ To compare biomarkers’ variability and temporal consistency, the coefficient of variation (CV%) of the three biomarkers across seven months of data at each of the 64 WWTPs was calculated. Only 7-months of data (n=1596) between the weeks of 09/13/2021 to 04/10/2022 were included in the analyses to compare three biomarkers since PMMoV was only measured after 09/13/2021. The relationship between weekly biomarker load and the population contributing to the wastewater was examined using linear regression. The biomarker load of biomarker *i* for *j* WWTP, *B*_*ij*_, was calculated as

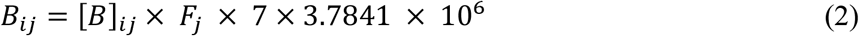

in which [*B*]_*ij*_, the biomarker *i* concentration in *j* WWTP wastewater sewershed, was determined by LC-MSMS or RT-qPCR. *F*_*j*_ is the wastewater daily flow volume (million gallons per day) for WWTP_*j*_. Constants 3.78541 and 7 are applied to convert the imperial unit to metric unit and daily to weekly biomarker load, respectively.

The linear regression model was conducted as

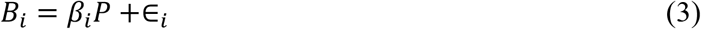

Where *P* is the corresponding population from metadata data. The modeling was based on the assumption that the mass load entering the treatment plant per day of a chemical is proportional to the contributing population. Since the actual real-time population was not available, the best estimation of it was the metadata population. *B*_*i*_ is the biomarker mass load, *∈*_*i*_ the error term, and *β*_*i*_ the estimated parameter for biomarker *i*. Log transformation was applied to the population and biomarker load.

In addition, ten wastewater samples (each from different WWTFs) from Wisconsin collected during the week of 06/07/2021 were used to validate equations (3) using Root Mean Square Error (RMSE). The RMSE is the standard deviation of the prediction errors.

Then, the real-time population *P*_*RT*_ by biomarker *i* was estimated using all three biomarkers according to the equation

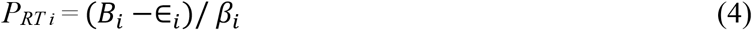

#### 2.5.2. Population normalization of wastewater SARS-CoV-2 load and COVID-19 incidence rate

Wastewater SARS-CoV-2 load was then normalized to population (copies/week/10,000 people) by dividing the SARS-CoV-2 load per week by each of the three estimated population metrics:

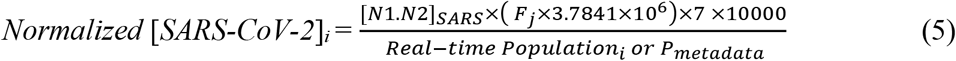

in which *[N1.N2]*_*SARS*_ (copies/L), the SARS-CoV-2 concentration in the wastewater sewershed, was determined by RT-qPCR using equation (1). The COVID-19 incidence rate was defined as:

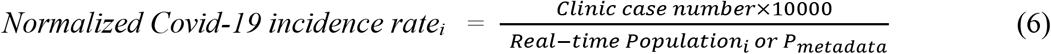

Population normalization better allows the results to be compared across WWTPs that serve different size populations.

#### 2.5.3. Effectiveness of population normalization on the relationships between wastewater SARS-CoV-2 load and clinical COVID-19 incidence rate

To compare the outcomes of population normalization of different biomarkers, the strength of the linear regression models from normalized wastewater SARS-CoV-2 load to normalized COVID-19 incidence rate were compared across markers, as well as when there was no population normalization (i.e., the correlation strength of raw viral load per week with total case number per week). Specifically, the goodness of fit of linear regression models was compared within each WWTP using the coefficient of determination (R^2^), the measure of “variance explained”. Analysis of variance (ANOVA) and pairwise comparisons were conducted on the R^2^ of four groups (without normalization, metadata population normalization, caffeine estimated population normalization, paraxanthine estimated population normalization, and PMMoV estimated population normalization) within each WWTP. Log transformation was applied to both wastewater SARS-CoV-2 load and the COVID-19 incidence rate. Since there are many “0” COVID-19 cases, log_10_ (x+1) was employed for the COVID-19 incidence rate.

## 3. RESULTS

### 3.1. Temporal variations of biomarkers

The coefficient of variations (CV%) of the three biomarkers across seasons at each of the 64 WWTPs was calculated (Fig. 2). The mean CV% across all the facilities were not different between caffeine (43%) and paraxanthine (40%), but both significantly lower than the CV% of PMMoV (mean=67%) (p<0.001). Paraxanthine was the only biomarker with all the CV% lower than 100% and the smallest range of CV% among 64 WWTPs (caffeine = 263%, paraxanthine = 54%, and PMMoV = 154%). The lower CV% means of caffeine and paraxanthine than PMMoV indicated that caffeine and paraxanthine changed less over time than PMMoV. No significant relationship between each biomarker’s CV% and population size was found (data not shown). Compared to caffeine and PMMoV, the smallest CV% range of paraxanthine among WWTPs showed paraxanthine is less influenced by population attributes (i.e., population size, the composition of age, race, and ethnicity, and environmental factors such as season and fecal strength across geographic regions).

**Fig. 2.**
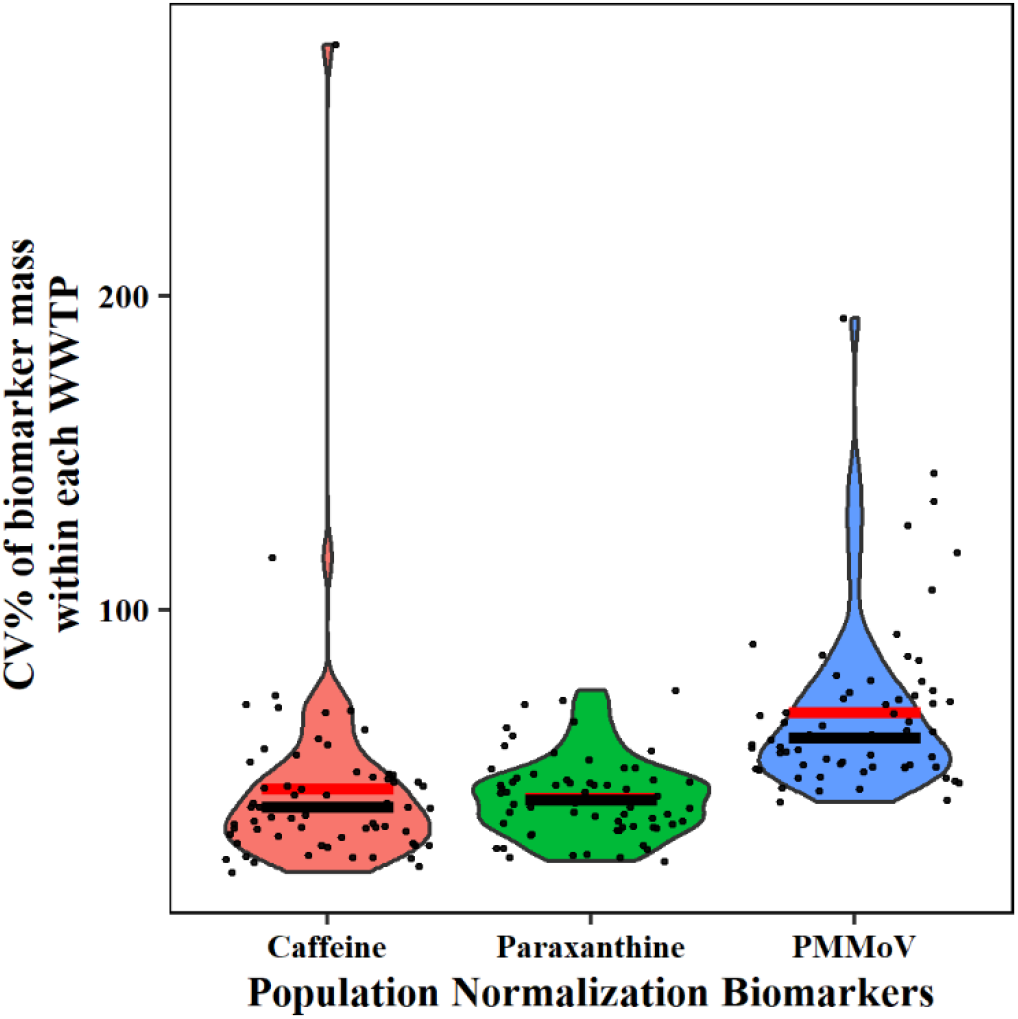
Distributions of coefficient of variation (CV%) of three biomarkers over seven months of study time from 09/13/2021 to 04/10/2022 (n=1569) within 64 WWTPs. The red lines indicate the mean CV%, and the black lines indicate the median CV% of each biomarker.

The outlier of CV% of caffeine (280%) was the Sikeston Wastewater Treatment Plant (SKSTN), which is nearby a global industry factory, Unilever ice cream, manufacturing the distributed worldwide Magnum chocolate ice cream bar (caffeine concentration in chocolate is around 420 μg/g). Eleven out of 16 samples with caffeine concentrations over 200 μg/L/week were from SKSTN. The extreme caffeine concentrations were not likely all shed by humans from such a small town with a population of 17,000 but rather from industrial waste. After removing samples from SKSTN, there was a strong linear relationship between caffeine and paraxanthine concentration (Fig. S1). Therefore, caffeine is still a reliable biomarker for the area where no such exogenous caffeine sources exist.

### 3.2. Real-time population prediction by biomarkers

Linear regression models were established for biomarker mass load per week and population sizes obtained from the metadata for both Missouri. Then, real-time population (*P*_*RT*_) was predicted for each WWTP with all available data. The estimated *P*_*RT*_ of tourism town (ANON2) from all three biomarkers was generally higher than the metadata population, which is reasonable since the metadata population primarily only counts residents. The estimated *P*_*RT*_ for the large metropolitan area, Kansas City Blue River, was almost always lower than the metadata population (Fig. S3A), probably indicating a population decline in the past two years since the census survey in 2020. There was a strong relationship between the metadata population and the total case number during the seven months from 09/13/2021 to 04/10/2022 (Fig. S3B and S3C). Kansas City Blue River was the “outlier” in Fig S3B, suggesting that the PRT of the Kansas City Blue River area might be substantially lower than the metadata population. Upon further investigation, it was learned that KCBLU WWTP was in the process of upgrading and the served area changed periodically compared to the metadata population that the facility initially provided (personal communication).

Therefore, models between biomarker mass load and population were revised by removing KCBLU WWTP in the models for Missouri (Fig. 3A, 3C, 3E) and metadata normalization data in KCBLU were also removed in the following analyses. Linear regression models were also established for biomarker mass load per week and population sizes obtained from the metadata for Wisconsin (Fig. 3B, 3D, 3F). Within Missouri, the model with paraxanthine had the highest R^2^ among all three biomarkers. Similar trends were observed in Wisconsin models, especially the paraxanthine model, which confirmed that the relationship between biomarkers and population could be applied beyond Missouri. In addition, Wisconsin samples were used to test Missouri models (Fig. 3A, 3C, 3E) using RMSE, which were 0.24, 0.19, and 0.87 for models of caffeine, paraxanthine, and PMMoV. The lowest RMSE of the paraxanthine model indicated it is the best predictor of the population too.

**Fig. 3.**
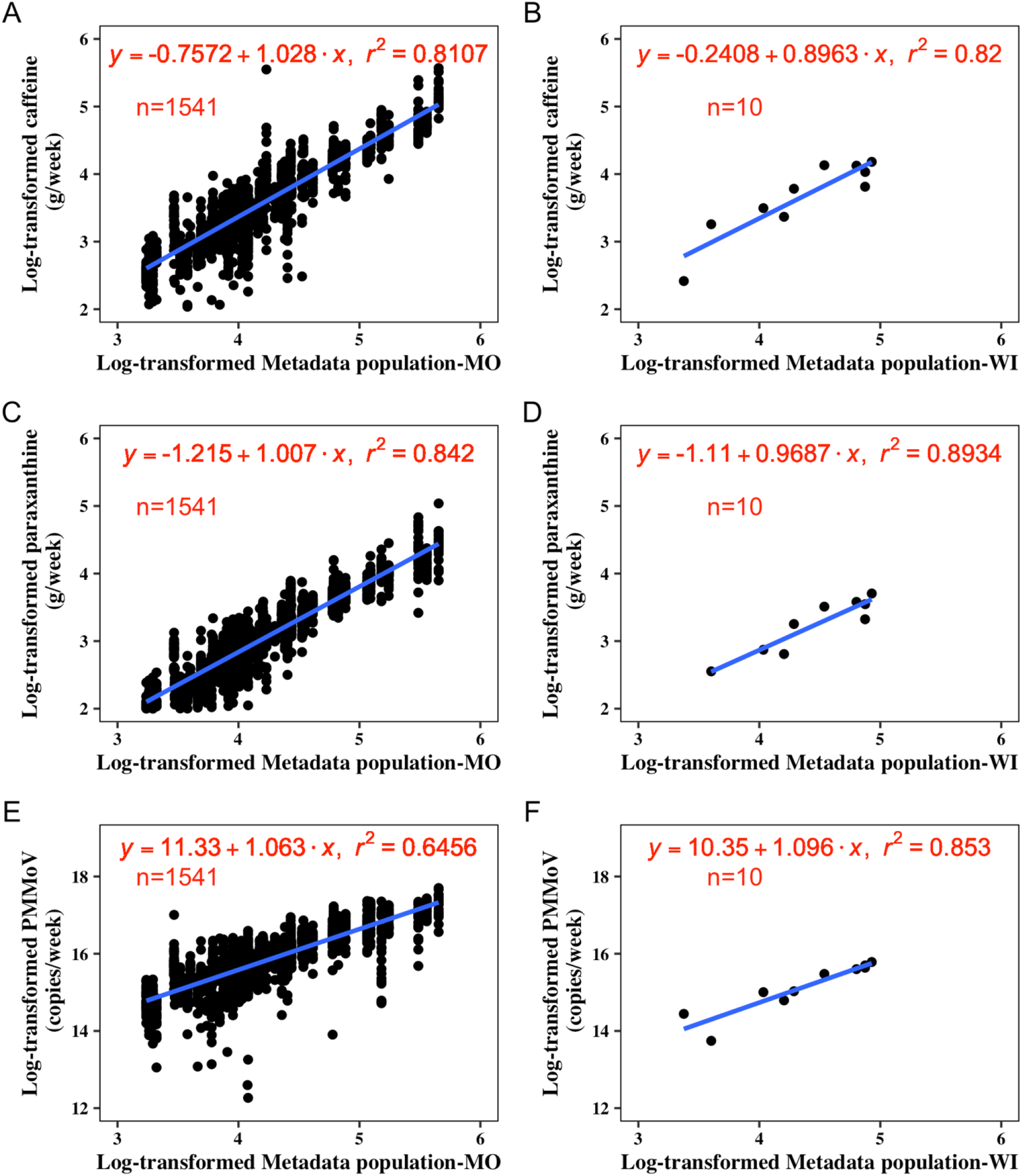
The linear regression models between the mass loads of three wastewater biomarkers [(caffeine, paraxanthine, and pepper mild mottle virus (PMMoV)] and Metadata population (GIS-mapped census population or estimated from sewer connections) for Missouri based on 7-month wastewater samples collected from 09/13/21 to 04/10/22 across 63 wastewater treatment plants (WWTPs) (plot A, C, and E). One facility from the initial 64 WWTPs (WWTP KCBLU) was removed from the models since the population served changed dramatically during the sampling period due to facility upgrading. Plots of B, D, and F were the models for Wisconsin based on ten samples collected during the week of 06/07/2021 from ten different WWTPs.

The ratios of the estimated real-time population to the metadata population (*P*_*RT*_/*P*_*METADATA*_) by paraxanthine had the smallest mean and standard deviation among the 64 WWTPs (Table 2). Among the three biomarkers, caffeine and paraxanthine, in general, are closer to the metadata populations. In contrast, estimated *P*_*RT*_ from PMMoV wavered with time dramatically. The smaller variations of estimated *P*_*RT*_ from caffeine and paraxanthine than PMMoV are consistent with the low temporal variations of caffeine and paraxanthine relative to PMMoV.

**Table 2.** Summary of ratios between estimated real-time population by three biomarkers to the Metadata population.

| **Ratios** | **Mean** | **Max** | **Min** | **SD** |
| --- | --- | --- | --- | --- |
| Caffeine-population/metadata population | 1.248 | 79.669 | 0.008 | 1.862 |
| Paraxanthine-population/metadata population | 1.137 | 12.053 | 0.008 | 0.741 |
| PMMoV-population/metadata population | 1.471 | 75.203 | 0.00008 | 2.26 |

A case study of validation of biomarker population estimation using the Apple mobility data was conducted in a college town, Columbia, Missouri. The Apple mobility showed that demographic migration was influenced by the COVID-19 pandemic and the school events in Columbia, Missouri (Fig. 4A). For instance, there was a sharp decline of mobility in April 2020 after the Missouri “Stay home” order. Both mobility data and the estimated *P*_*RT*_ estimated by paraxanthine showed the fluctuations of the population with school semesters and holidays (Fig. 4A and B). For instance, Labor Day weekend, Thanksgiving break, and winter break had lower *P*_*RT*_ and mobility than other times since a large group of students traveled back to their hometowns during these holidays. Also, the week of the University of Missouri Homecoming event (10/05/2021) had a *P*_*RT*_ and mobility increase.

**Fig. 4.**
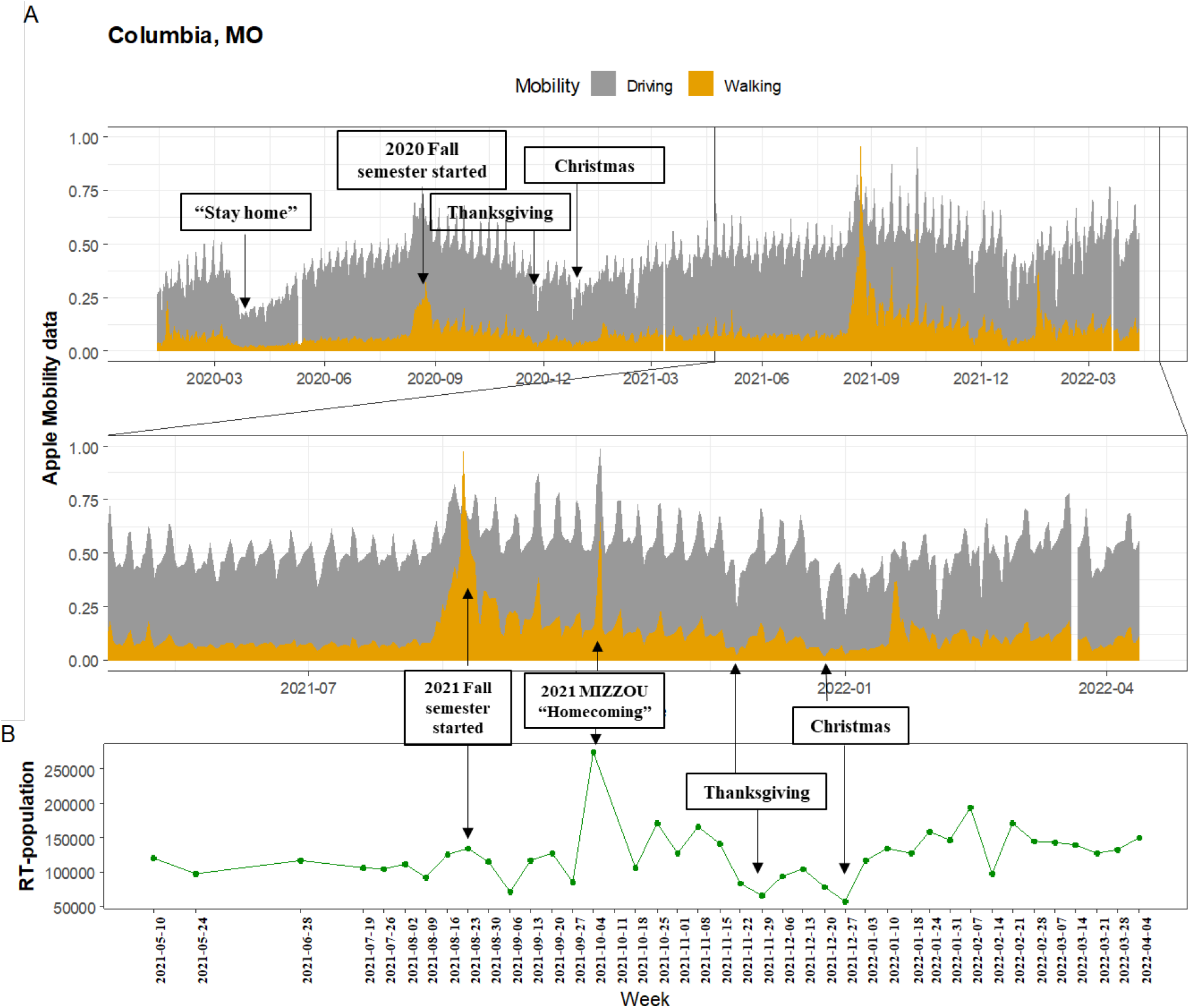
Apple mobility indices of Columbia, MO for walking and driving using aggregated data collected from Apple Maps data for the duration of 01/13/2020 to 04/10/2022 (A). The original Apple mobility indices have been scaled to the maximum observed during the study period. Predicted real-time population using paraxanthine in Columbia, MO (B).

### 3.3. Effectiveness of population normalization of different wastewater biomarkers

The strengths of the relationships between the population normalized SARS-CoV-2 RNA load and COVID-19 incidence rate were compared within each WWTP (Table 3). A total of 59 out of 64 WWTPs showed a significant positive relationship between wastewater SARS-CoV-2 RNA load and clinical COVID-19 incidence rate for all population normalization scenarios. In addition, models were significant except for the PMMoV estimated population normalization model for two WWTPs (ANON1 and MACON). Within 57 of 64 WWTPs, biomarker normalizations strengthened the relationship between SARS-CoV-2 RNA load and normalized COVID-19 incidence rate (i.e., R^2^ increased). The mean R^2^ decreased (α level=0.1) in the order: paraxanthine estimated real-time population normalization > without population normalization and metadata population normalization > caffeine estimated real-time population normalization > PMMoV estimated real-time population normalization, indicating that paraxanthine is the best population normalization biomarker (Fig. S3).

**Table 3.**
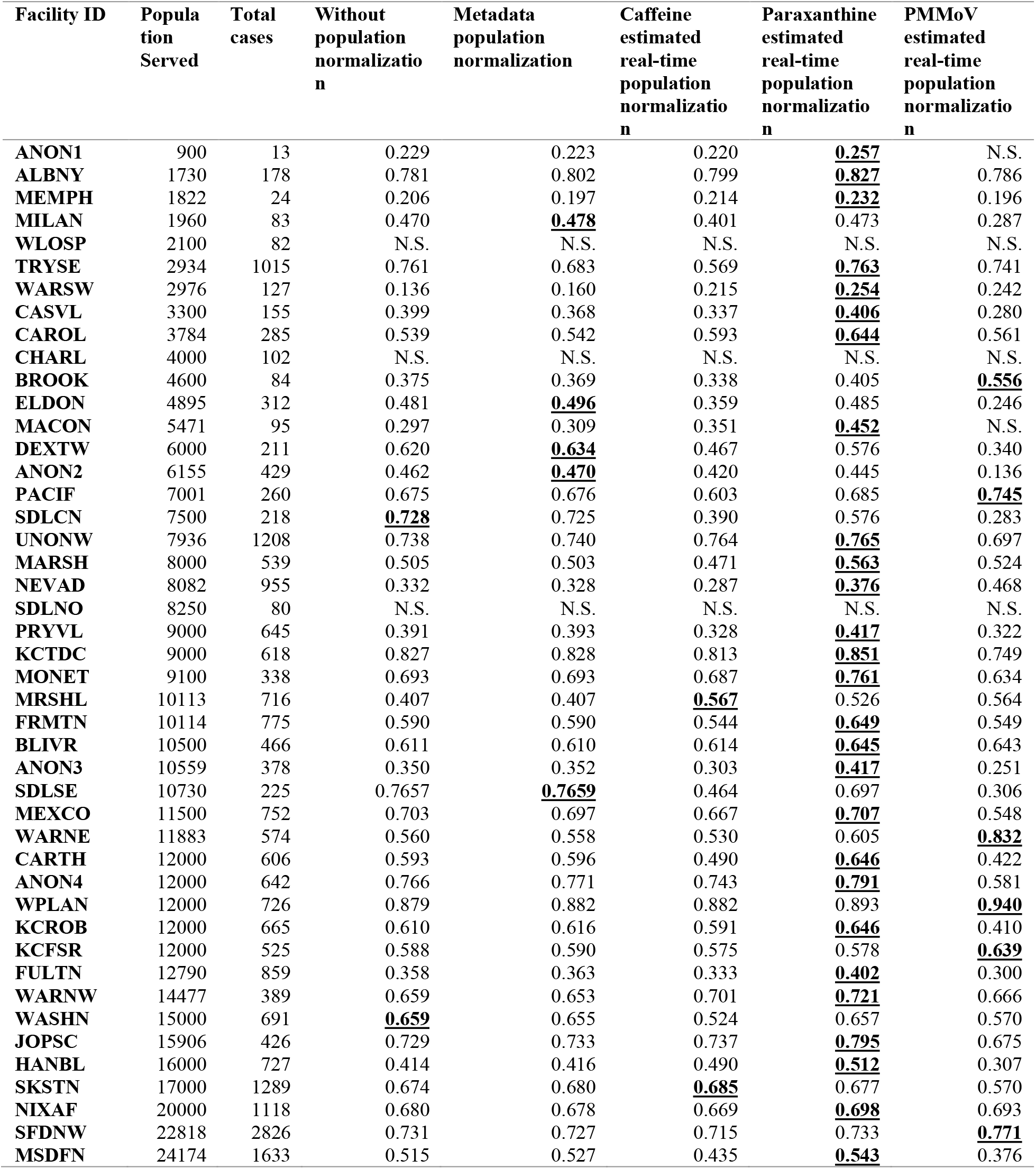
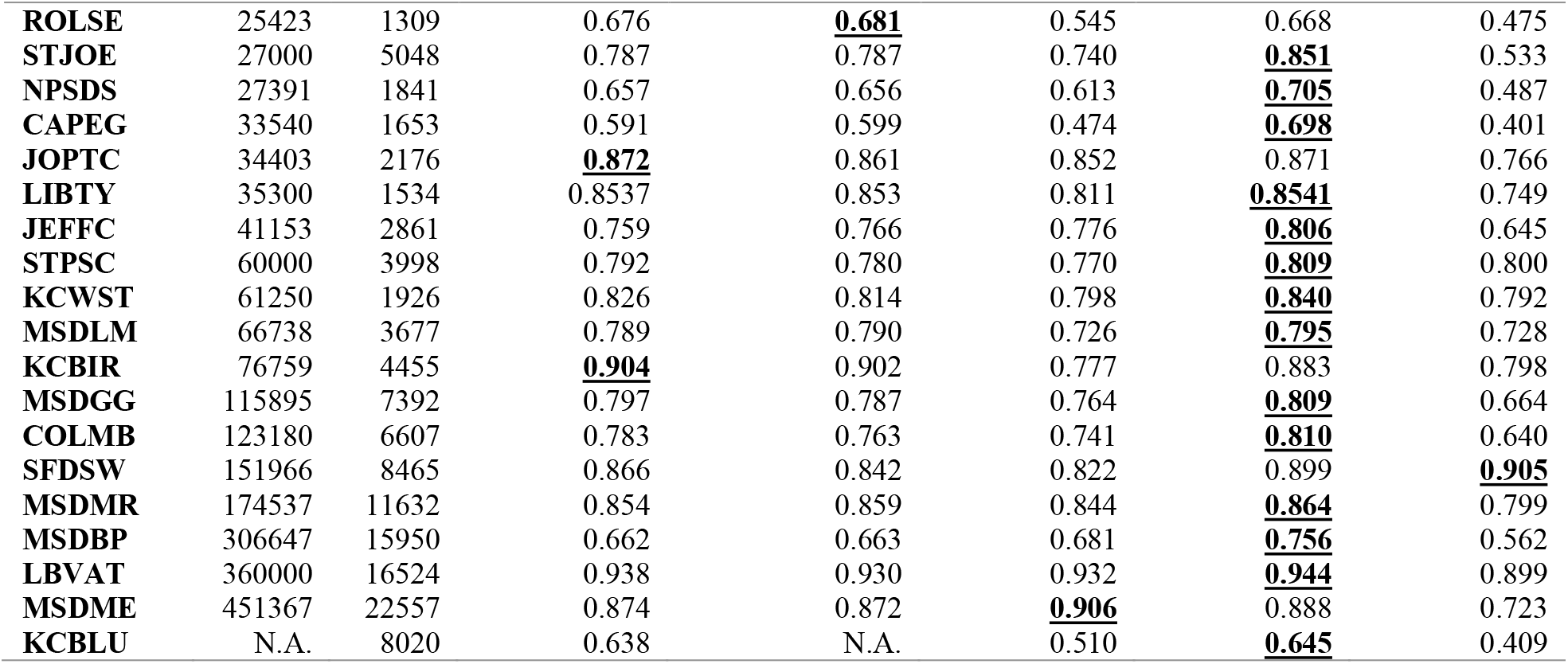
Coefficient of determinations (R^2^) of the linear regression models between log-transformed wastewater SARS-CoV-2 RNA load (copies/week/10K person) and clinical COVID-19 incidence rate (case number/week/10K person) within each WWTP without and with population normalization by metadata population, and the real-time populations estimated from caffeine, paraxanthine, and PMMoV. The bold and underlined values are the highest R^2^ for each WWTP among five models. “N.S.” indicates the not significant models. “N.A. indicated not available information.

The regression models of viral loads with clinical incidence after normalization to paraxanthine estimated population had the highest R^2^ among all five models for 40 out of 61 WWTPs (Fig. 5). In contrast, the strength of the relationship between wastewater viral load and clinical incidence after PMMoV and caffeine estimated population normalization became weaker than without population normalization or with metadata population normalization (Fig. S3). Therefore, the time series of wastewater viral load and COVID-19 incidence rate with normalization of paraxanthine estimated population was plotted for each WWTP (Fig. 6). Weekly wastewater viral copies followed similar patterns with clinical incidence rates for most WWTPs. Furthermore, the strength of the relationship between wastewater SARS-CoV-2 viral load and the COVID-19 incidence rate was significantly increased with increasing cases and metadata population (Fig. 7A and 7B).

**Fig. 5.**
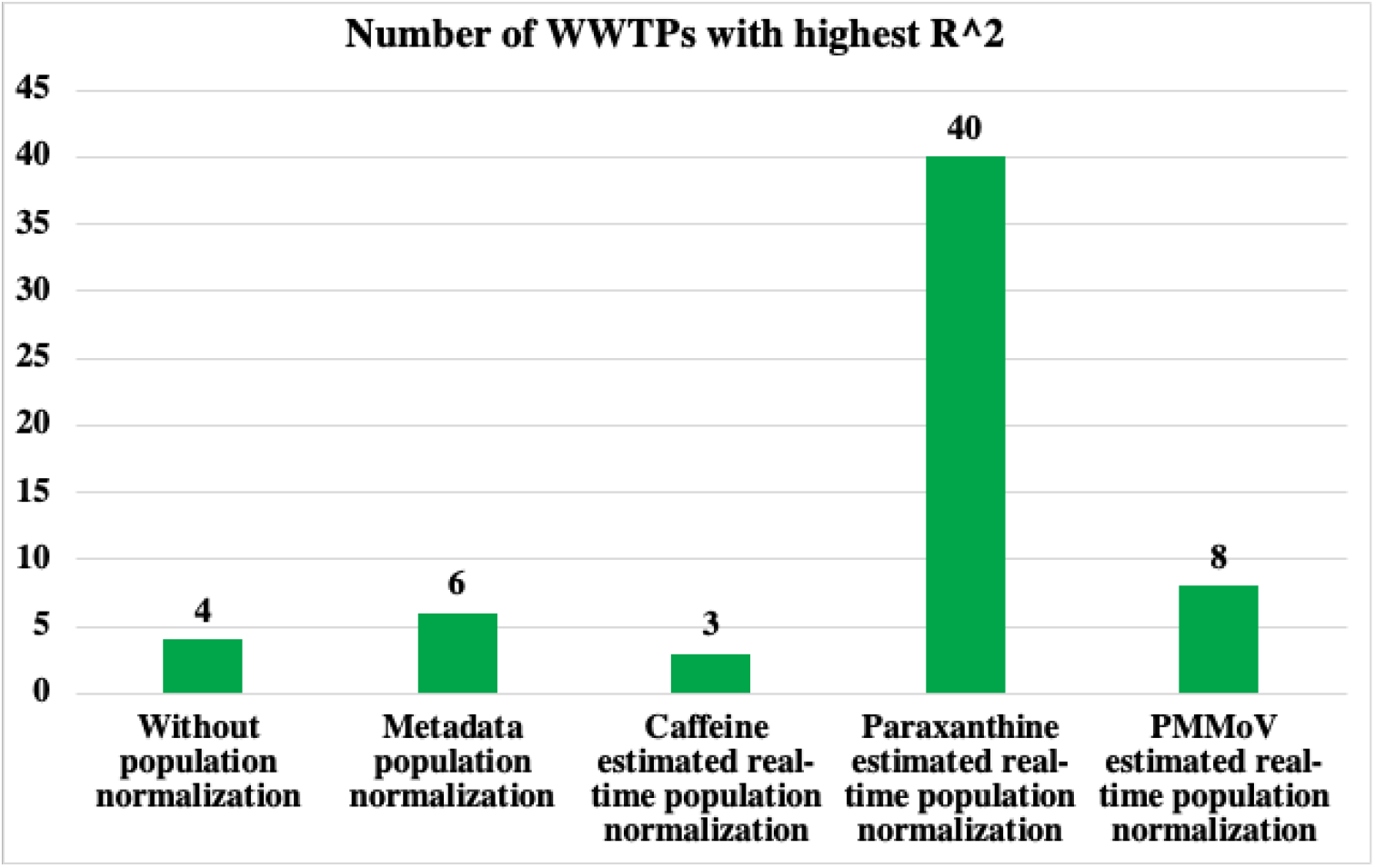
The WWTP numbers with best population normalization models (highest R^2^) for each normalization model. The “61” after slash indicated the 61 out of 64 WWTPs had significant linear regression models between wastewater SARS-CoV-2 RNA load (copies/week/10K person) and clinical COVID-19 incidence rate (case/week/10K person). The numbers before the slash indicated the number of WWTPs with the highest R^2^ among the five regression models.

**Fig. 6.**
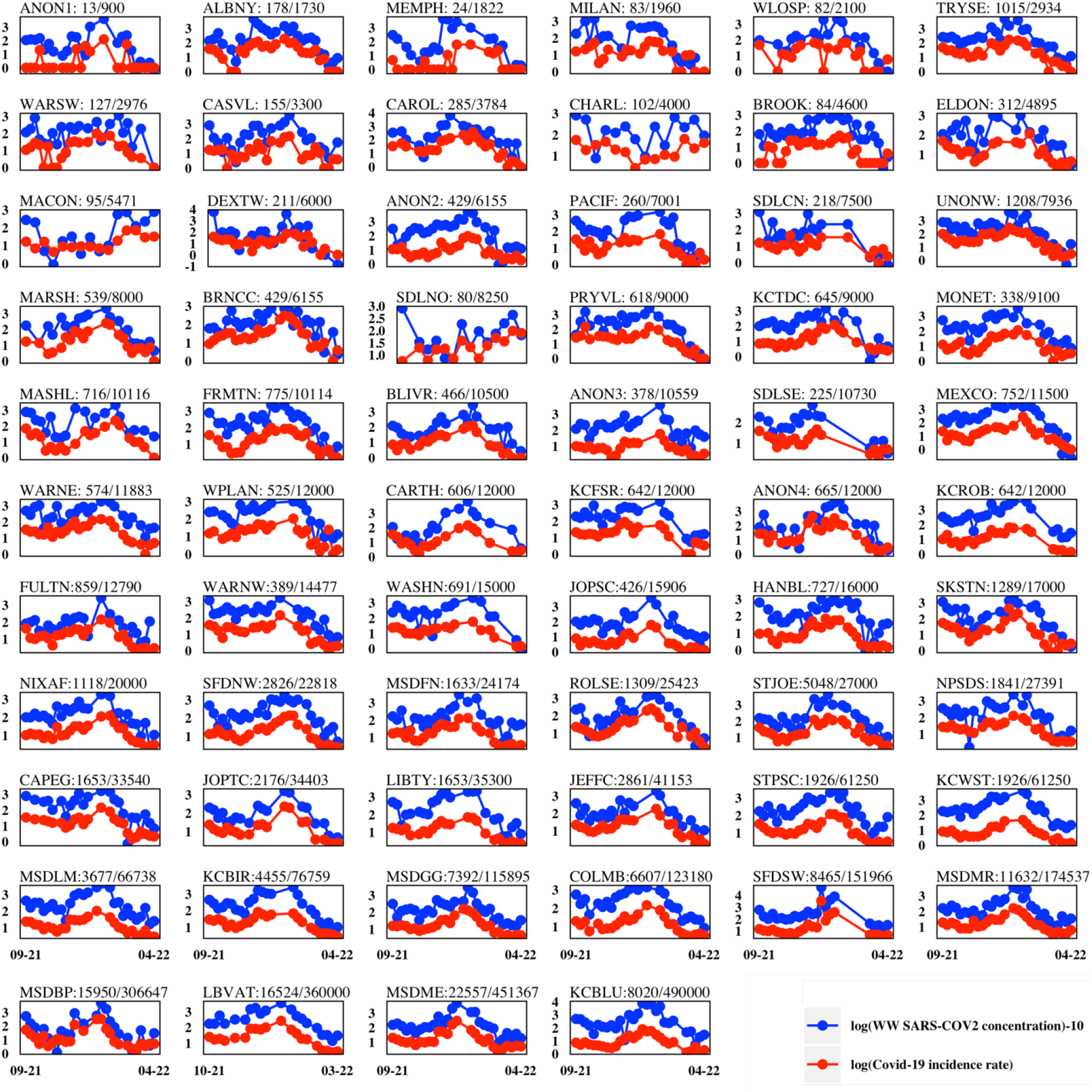
Normalized wastewater SARS-CoV-2 RNA load (copies/week/10K person) by paraxanthine estimated real-time population and clinical COVID-19 incidence rate (case/week/10K person) within 64 WWTPs from the weeks of 09/13/2021 to 04/05/2022. The population of WWTPs increase from top left to right down. The title of each plot is in the format of “WWTP name: total COVID-19 case number/Metadata population”.

**Fig. 7.**
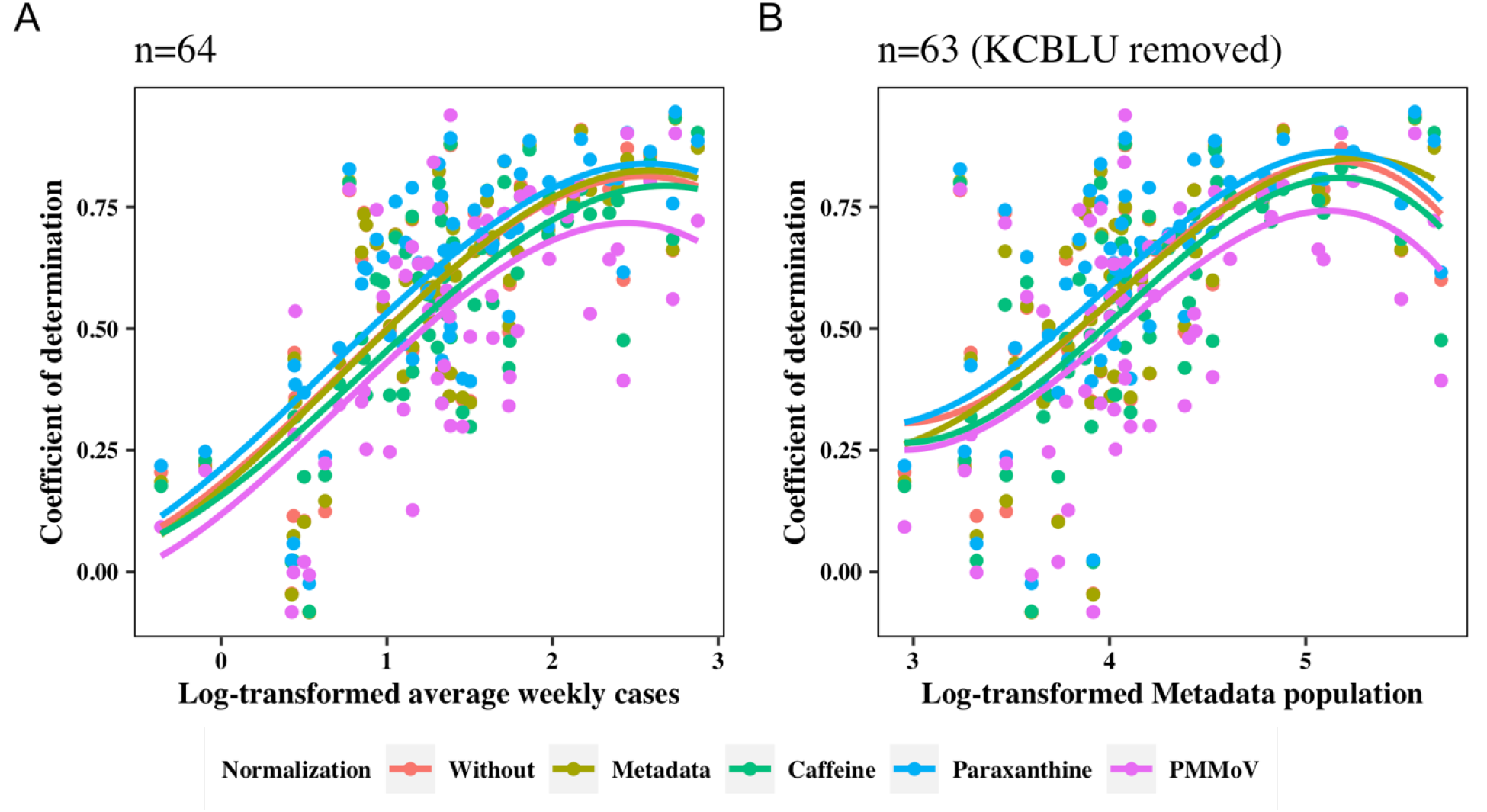
The polynomial relationships between the coefficient of determination (R^2^) of the linear regression models and clinical case (A) and Metadata population (B). The models were linear regressions between wastewater SARS-CoV-2 RNA load (copies/week/10K person) and clinic COVID-19 incidence rate (cases/week/10K person) within 64 WWTPs without and with the population normalizations.

### 3.4. Limit of detection of wastewater SARS-CoV-2 surveillance

The linear regression relationship between wastewater SARS-CoV-2 RNA load with clinical cases was tested for models without population normalization and paraxanthine population normalization (Fig. S4). According to the paraxanthine population normalization model, the process limit of detection (PLOD) (entire process from the sampling to the analysis of RT-qPCR) in the wastewater over should be:

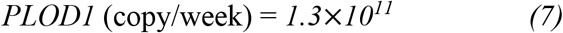

A validation calculation was conducted using the PLOD (<3,954 GC/50ml; 95% probability of detection) of the US CDC N1 RT-dPCR and RT-qPCR assays from a study based on wastewater SARS-CoV-2 seeding experiment by Ahmed et al.^34^ (Table 4):

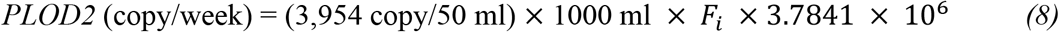

**Table 4.** Case study in the smallest WWTP ANON1 (900 population) for the process limit of detection of wastewater SARS-CoV-2 surveillance. The process limit of detection (PLOD) of N1N2 mass load PLOD1 for at least one case was calculated based on the results of this study (Figure 7B). The PLOD2 was calculated based on the literature.^34^ The bold and underling values were the three weeks that N1N2 mass load higher both PLOD1 and PLOD2.

| SWCTY Cases | Flow rate (GMD) | Week | N1N2 mass load | N1N2 mass - PLOD1 | N1N2 mass - PLOD2 |
| --- | --- | --- | --- | --- | --- |
| 0 | 0.0425 | 9/13/2021 | 5.56E+10 | -7.44E+10 | -3.35E+10 |
| 0 | 0.0639 | 9/20/2021 | 7.10E+10 | -5.90E+10 | -6.29E+10 |
| 0 | 0.0535 | 9/27/2021 | 9.22E+10 | -3.78E+10 | -1.99E+10 |
| 1 | 0.0531 | 10/04/2021 | 7.85E+10 | -5.15E+10 | -3.28E+10 |
| 0 | 0.0553 | 10/11/2021 | 1.62E+10 | -1.14E+11 | -9.97E+10 |
| 0 | 0.0488 | 10/18/2021 | 4.92E+10 | -8.08E+10 | -5.31E+10 |
| 0 | 0.0562 | 10/25/2021 | 1.38E+10 | -1.16E+11 | -1.04E+11 |
| 0 | 0.0546 | 11/8/2021 | 6.63E+09 | -1.23E+11 | -1.08E+11 |
| 0 | 0.0512 | 11/15/2021 | 1.45E+10 | -1.16E+11 | -9.28E+10 |
| 0 | 0.0474 | 11/22/2021 | 8.18E+10 | -4.82E+10 | -1.75E+10 |
| 1 | 0.0458 | 11/29/2021 | 7.91E+10 | -5.09E+10 | -1.69E+10 |
| 0 | 0.0493 | 12/6/2021 | 1.54E+10 | -1.15E+11 | -8.79E+10 |
| 2 | 0.053 | 12/13/2021 | 4.32E+09 | -1.26E+11 | -1.07E+11 |
| **<u>1</u>** | **<u>0.0544</u>** | **<u>12/20/2021</u>** | **<u>7.34E+11</u>** | **<u>6.04E+11</u>** | **<u>6.20E+11</u>** |
| **<u>6</u>** | **<u>0.0544</u>** | **<u>1/10/2022</u>** | **<u>1.98E+12</u>** | **<u>1.85E+12</u>** | **<u>1.87E+12</u>** |
| 0 | 0.0573 | 1/31/2022 | 6.01E+10 | -6.99E+10 | -6.00E+10 |
| **<u>0</u>** | **<u>0.06</u>** | **<u>2/7/2022</u>** | **<u>1.37E+11</u>** | **<u>7.00E+09</u>** | **<u>1.13E+10</u>** |
| 2 | 0.06 | 2/14/2022 | 9.48E+10 | -3.52E+10 | -3.09E+10 |
| 0 | 0.0608 | 2/21/2022 | 4.69E+09 | -1.25E+11 | -1.23E+11 |
| 0 | 0.06 | 2/28/2022 | 3.36E+08 | -1.30E+11 | -1.25E+11 |
| 0 | 0.072 | 3/7/2022 | 1.44E+09 | -1.29E+11 | -1.49E+11 |
| 0 | 0.07 | 3/14/2022 | 9.97E+08 | -1.29E+11 | -1.46E+11 |
| 0 | 0.11 | 3/21/2022 | 2.18E+09 | -1.28E+11 | -2.28E+11 |
| 0 | 0.05 | 3/28/2022 | 6.19E+08 | -1.29E+11 | -1.04E+11 |

Although Ahmed et al. used a different adsorption extraction method of virus to evaluate N1 copy instead of average copy of N1 and N2 as in this study, both PLOD1 and PLOD2 showed that the N1N2 mass load was higher than PLODs for ANON1 (the smallest WWTP) during the three weeks when the main outbreak occurred. The PLOD1 was likely higher than the actual value since the clinical cases are likely less than the real cases due to the asymptomatic and underreported cases.

## 4. DISCUSSION

### 4.1 Paraxanthine was the optimal population biomarker

The reduced strength of the relationship between the PMMoV-population normalized SARS-CoV-2 load and COVID-19 incidence rate was consistent with previous studies that showed PMMoV had mixed or adverse effects on the correlation between wastewater measurements of SARS-CoV-2 and clinical cases.^35–37^ To the best of our knowledge, there is only one nationwide study to date (conducted by Biobot Analytics) that is comparable with our study in terms of the temporal and spatial magnitudes of sampling. That study collected 2,433 samples from 55 WWTPs across the U.S. and found that PMMoV normalization did not always improve the correlation between wastewater measurement and clinical cases.^35^ Feng et al. showed that PMMoV normalizations reduced the correlations between SARS-CoV-2 concentration and COVID-19 incidence for 8 of 12 WWTPs and suggested that variability’s influence across measurement for human viral is stronger than that of differences in the fecal loads in the samples^36^. For some sewersheds where the normalizations by metadata population and without population normalization were better than the normalizations by caffeine and PMMoV estimated population, probably because most WWTPs serve rural areas in Missouri where the population does not fluctuate with time. However, each WWTP was weighted equally in this analysis regardless of the population size it serves. Furthermore, many people worked from home during the pandemic; therefore, the real-time population was expected to be closer than usual to the metadata population for many regions.

The intensified relationship between wastewater SARS-CoV-2 viral load with clinical COVID-19 cases by wastewater paraxanthine concentration for 2/3 of the WWTPs and the consistency of the relationship between wastewater paraxanthine concentration and population between Missouri and Wisconsin demonstrated that paraxanthine is a reliable population biomarker across large geographical regions with different sizes of the population. The superior performance of paraxanthine over PMMoV could be attributed to its 1) much longer half-life, 2) less exogenous sources and variability of excretion rate intra-individual and inter-individual, 3) easier and more accurately determined in the wastewater.

#### 4.1.1 Stability

A promising population normalization biomarker should be stable in wastewater. Caffeine and its metabolites in untreated wastewater were stable during 24 hr storage at 4°C and 20°C and also stable at −20°C for up to four weeks.^28^ One study showed PMMoV was stable for 21 days at 4°C, 25°C, and 37°C,^38^ However, it was mostly based on different water types that had simpler contents than wastewater, such as deionized water and autoclaved wetland water. In addition, the conclusion that PMMoV has high stability over time was drawn from much shorter periods of investigations or a small number of samples collected from fewer WWTPs than in our study^4,18,21,22,36^. For instance, a highly cited article from Kitajima et al. found that PMMoV did not change seasonally, but was only based on 48 samples collected from two WWTPs over 12 months.^21^ Wu et al. only tested PMMoV stability for SARS-CoV-2 fecal biomarker on samples collected over one week during March 2020.^4^ D’Aoust et al. found that PMMoV is a better normalization biomarker than *Bacteroides* 16S rRNA or human 18S rRNA, which was also only based on 23 samples from two WWTPs.^18^ In contrast, higher stability of paraxanthine than PMMoV in our study was based on 1569 samples from September 2021 to April 2022, which covered three months of the Delta variant in 2021 and the major spike of the Omicron variant at the beginning of 2022. The 64 WWTPs represent 50% of the entire Missouri population, from metropolitan areas such as St. Louis to small rural towns with only 900 people.

#### 4.1.2. Sources

A reliable population biomarker should primarily be shed by humans and respond to the population size, not environmental factors.^8^ A regular and constant consumption is a further prerequisite for a good marker.^39^ Loads of caffeine in untreated wastewater reflect not only consumption, metabolism, and excretion of the compound but also caffeine from beverages, foods, and pharmacologic that were poured out directly.^40^ Caffeine is transformed in the human liver into more than 20 metabolites, primarily dimethylxanthines (paraxanthine, theobromine, and theophylline), dimethyl- and monomethyluric acids, and uracil derivatives^39^. Between 0.5% and 10% of the caffeine in human body is excreted unmetabolized via urine.^39^ Exogenous sources from industrial wastewater can influence caffeine if the WWTP is a combined sewer system, which also has potential pollution issues from the outflow.^7^ Besides, we also had several samples that could not detect caffeine and paraxanthine after heavy storms (data removed from analyses). Therefore, if combined sewer systems are chosen for WWTP, individuals should be aware that in some circumstances, like high rain events, readings of viral load and other measurements may need be corrected for dilution.

As a caffeine metabolite, paraxanthine mainly comes from human metabolism.^8^ Therefore, paraxanthine relates to the population better than caffeine. Among the metabolites of caffeine, paraxanthine is the most abundant caffeine metabolite in wastewater.^26^ In addition, 1-methylxanthine, 7-methylxanthine, and 1,7dimethyluric acid are also metabolites of theophylline and theobromine respectively, which are present in some foods, drinks, and pharmaceutical formulations.^39^ Paraxanthine was believed to be the optimal biomarker of caffeine intake as a population biomarker.^1^ The more minor variations of paraxanthine among different WWTPs were probably due to similar levels of caffeine intake among different groups in the population. The main factor driving paraxanthine load in the wastewater was the coffee consumption rate, which can be influenced by the composition of the population age since kids do not drink as much as adults. In addition, average consumption is 70 mg per person per day but varies in the different countries due to the different culture.^39^ However, caffeine is one of the most widely consumed dietary ingredients worldwide, and thus paraxanthine as its metabolite has the potential to be used as a population biomarker worldwide.

In comparison, PMMoV is a pepper pathogen virus that often is found in human feces, as well as peppers and processed pepper products from all over the world, such as dry spices and sauces.^42^ Unlike caffeine or tea are widely consumed by people all over the world, the importance of pepper in cuisine varies depending on the regions. For example, chili pepper is the defining ingredient of New Mexican food but not for most European cuisine. Therefore, the population race composition might influence the pepper’s consumption and thus PMMoV concentration. The detection level of PMMoV in human feces varies greatly from 7% to 95%, depending on the study’s regions and also between adults and children, even in the same regions.^43^ The PMMoV concentration in our study had an average of 1.5 *10^8^ copy/L, which is consistent with the previous study that showed the PMMoV concentration from raw wastewater ranged from 10^8^-10^10^ copy/L,^44^ but PMMoV in our study largely ranged from 3*10^4^ to 2*10^9^ copy/L.

In addition, SARS-CoV-2 RNA and PMMoV are shed in fecal, but caffeine and paraxanthine are discharged through urine. Humans urinate much more frequently than bowl movement, likely contributing to less variance in quantity among individuals than bowel movement, which possibly is one of the reasons that biomarkers in urine such as caffeine and paraxanthine had less variations and are more representative of population size.

#### 4.1.3. Quantification

Being easy to be determined with high repeatability is another criterion of a good population marker. The low variation of caffeine and paraxanthine owes to the high analytical sensitivity of LC-MSMS system and the consistent and high extraction recovery rate. With the development of technology, most of LC-MSMS systems have ng/mL to pg/mL level sensitivity. The instrumental detection limits for caffeine and paraxanthine were reported as 1.4 and 2.4 pg/injected, and instrumental quantification limits for caffeine and paraxanthine were 3.6 and 6.6 ng/L using API5500 QqQ equipped with a Turbo Ion Spray source.^28^ The sensitivity of our HPCL-MSMS with electrospray ionization is at ng/mL, but it is sufficient for wastewater caffeine and paraxanthine quantification. The caffeine concentration in wastewater influent water was reported from 20-300 μg/L (Canada), 20 μg/L (U.S.), and 147±76 μg/L (Germany).^28^ In our study, the average caffeine and paraxanthine concentrations were 71 ug/L and 17.5 ug/L, respectively. The recoveries of caffeine and paraxanthine from untreated wastewater were 88% and 76% during the similar storage temperature and extraction method to our study.^28^ Our previous study showed that the recovery rates of caffeine and paraxanthine during injection are 101±7% and 92±3%.^29^ In addition, the repeatability for caffeine and paraxanthine was quite high (CV%: 12% and 5%).^28^

Unlike the simple chemical analysis on LC-MSMS, PMMoV measurement has a very complex workflow. First, wastewater samples require the application of concentration steps before extraction of the RNA fragments. Then, highly-sensitive molecular assays using RT-qPCR or RT-dPCR (digital PCR) will be applied to quantify the PMMoV concentration.^34^ Consequently, there are many factors that may contribute to the large variation, such as the efficiency of primary concentration, loss through nucleic acid extraction, and inhibition of reverse transcription or PCR amplification. PMMoV’s recovery rates reported in previous studies varied but were generally lower than caffeine and paraxanthine (e.g., 45±26% using direct extraction and only >10% using electronegative (H.A.) filters).^36,45^ The recovery rate of PMMoV in this study was not tested directly, but the virus concentration methods (PEG concentration) in this study preserved SARS-CoV-2 N1N2 at approximately 62% signal on average and 2.7 times higher for Puro.^32^ The PMMoV’s process limits of quantification and detection were evaluated in diluted wastewater in the coastal water,^46^ which is different from PMMoV in raw wastewater.

### 4.2 Normalization approach recommendation for SARS-CoV-2 wastewater-based epidemiology

A previous study showed that outbreaks could be detected in buildings with as many as 1,000 occupants.^47^ This study showed that the wastewater could also detect community-level outbreaks with a small population size, which revealed that the process limit of detection for wastewater surveillance (e.g., the fewest infections in a community that can be reliably detected in wastewater) is quite low (∼0.1% of the population, 1-2 cases in 900) (Table 4). However, the PLOD could be overestimated since the actual cases could be higher than just 1 or 2 cases. The smallest WWTP, ANON1, still showed a significant relationship between the wastewater viral copies and the COVID-19 incidence rate, except when using the normalization of PMMoV estimated population (Table 3). Our previous study found that approximately 20% of the tested WWTPs in Missouri, U.S., receive some input from industries, possibly discharging some chemicals that suppress the viral signals in wastewater.^48^ MACON WWTP was one of the examples.^48^ However, positive relationship between the wastewater viral load and the COVID-19 incidence rate for MACON except for PMMoV estimated population normalization (Table 3). Therefore, WBE is a feasible approach for population-level monitoring of COVID-19 disease. PMMoV, however, is not an ideal population biomarker, especially for small towns due to its large temporal variation.

Many wastewater SARS-CoV-2 surveillance studies have been conducted across the U.S. during the last two years^35^, which provided an excellent network for the effective and long-term monitoring of SARS-CoV-2 and possibly other diseases in the future. Thus far, however, the normalization applied to the SARS-CoV-2 wastewater surveillance does not have a standardized approach. There are three main types of normalization: 1) normalized to WWTP flow (e.g., copies/week, to give viral load), 2) normalized to WWTP human fecal biomarker estimated population (e.g., copies/10K people/week), and 3) directly normalized to WWTP human fecal biomarker loads (e.g., copies/copy of PMMoV/week).

Wastewater flow normalization converts the measured viral concentration to viral load, accounting for variations in flow between days due to precipitation, snowmelt, or groundwater inflow. Established on the flow normalization, the normalization to biomarker estimated population evaluated in this study aims to account for the variations caused by wastewater volume and population size that contribute to the waste. The third normalization is used often because it does not involve wastewater volume and population information. However, it is based on the assumption that the measured wastewater biomarker concentration perfectly reflects the population dynamics. Many previous studies used PMMoV as SARS-CoV-2 internal control to normalize SARS-CoV-2 concentration to SARS-CoV-2 copies per copy of PMMoV.^4,18,49^ However, our study showed that the relationship between PMMoV and population is not as stable over time as caffeine and paraxanthine. In addition, our previous study confirmed that direct normalization effects of SARS-CoV-2 concentration using biomarker concentrations were always less ideal than indirect normalization, which involved the wastewater volume.^29^

Our findings suggest that for long-term wastewater SARS-CoV-2 surveillance, normalizing SARS-CoV-2 wastewater concentrations with a reliable population marker prior to calculating trends is highly recommended to account for changes in wastewater dilution and differences in relative human waste input over time due to tourism, weekday commuters, temporary workers, and pandemic lockdowns, etc. This approach is particularly critical for the sewershed with dynamic population changes, such as colleges, tourist towns, and metropolitan areas, where a large number of commuters who used to travel to cities daily transitioned to fully or partially remote workers after the pandemic. The relation between 1) size of the population, and 2) strength of the relationship between wastewater SARS-CoV-2 viral concentration and the COVID-19 clinical incidence rate was first demonstrated in this study. This would provide an excellent selection criterion for site selection, surveillance planning and data interpretation for the SARS-CoV-2 and even other wastewater-based epidemiology.

## 5. CONCLUSIONS

This study compared the effectiveness of three wastewater biomarkers for population normalization in the SARS-CoV-2 wastewater-based epidemiology with a large number of wastewater facilities across Missouri and long-term sampling over seven months. We found that PMMoV, one of the widely used population biomarkers for SARS-CoV-2 wastewater-based epidemiology, is not an ideal population biomarker since it reduced the strength of wastewater SARS-CoV-2 viral load and the COVID-19 incidence rate compared to the metadata population and without population normalization. Instead, paraxanthine, with many benefits, such as high stability, low exogenous sources than its precursor (caffeine), higher recovery rate, and low quantification variation, is very promising for predicting the real-time population and population normalization in the wastewater SARS-CoV-2 surveillance study, no matter the size of the population. The utility of this promising biomarkers was validated by data from ten different Wisconsin’s WWTPs with gradients in population sizes. The estimated real-time population using this biomarker was directly compared against the population patterns with human movement mobility data. Of the three biomarkers, population normalization by paraxanthine significantly strengthened the relationship between wastewater SARS-CoV-2 viral load and COVID-19 incidence rate the most (40 out of 61 sewersheds). Our findings suggest that paraxanthine has the potential to be serve as real-time population biomarker in other scenarios beyond SARS-CoV-2 wastewater-based epidemiology and not limited within Missouri geographical boundary.

## Data Availability

All data produced in the present study are available upon reasonable request to the authors

## 6. ACKNOWLEDGEMENT

The authors would like to thank the Missouri Department of Health and Senior Services (DHSS) for administrating the funding. We would like to express our gratitude to the Missouri Department of Natural Resources (DNR) for coordinating the sample collection. Research reported in this publication was supported by funding from the Centers for Disease Control and the National Institute on Drug Abuse of the National Institutes of Health under award number U01DA053893-01. We would also like to thank the Center for Agroforestry at the University of Missouri, USDA/ARS Dale Bumpers Small Farm Research Center under agreement number 58-6020-6-001 from the USDA Agricultural Research Service for supporting part of this research. The content is solely the responsibility of the authors and does not necessarily represent the official views of the National Institutes of Health, the Centers for Disease Control or USDA-ARS.

---

**Table S1.**
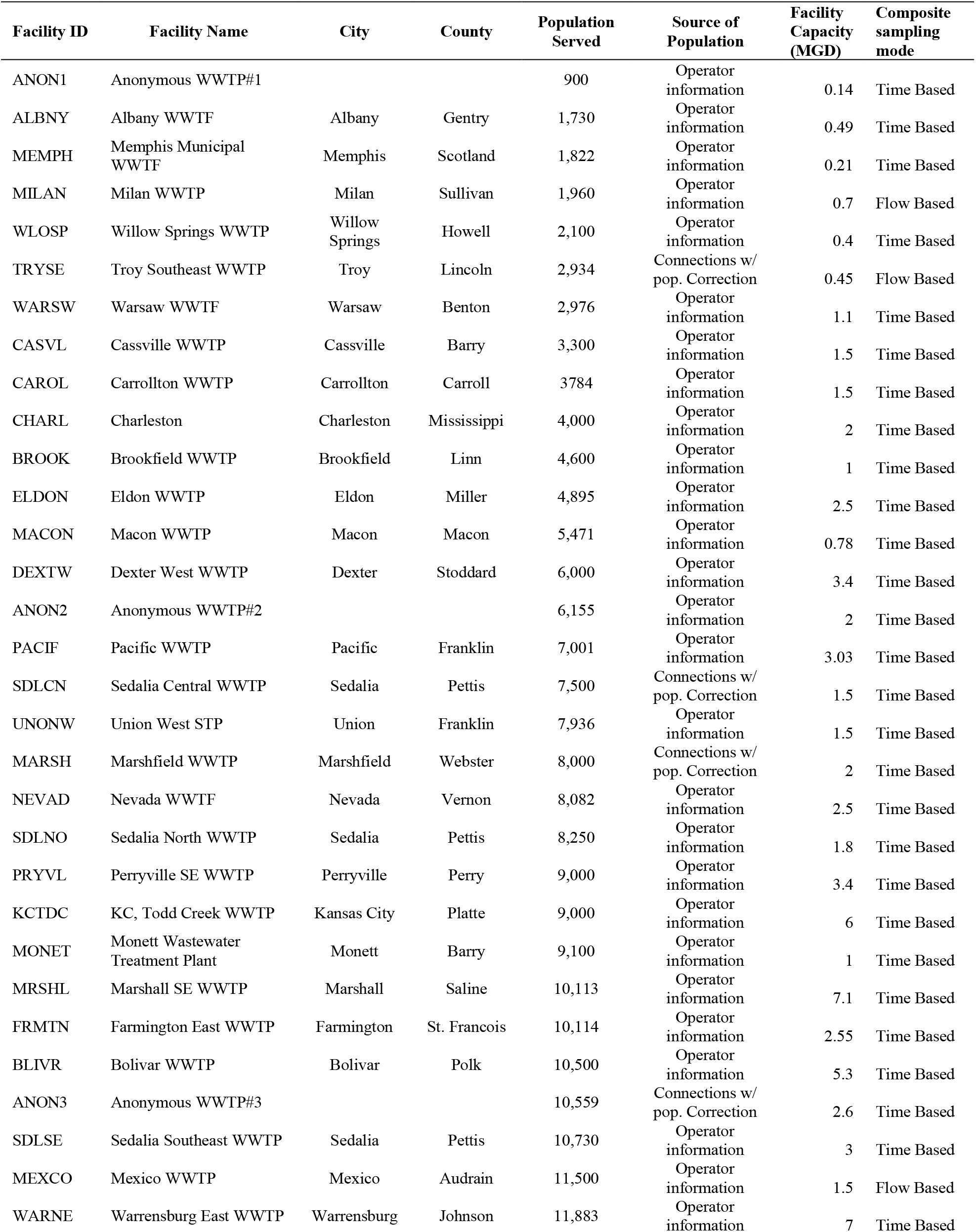
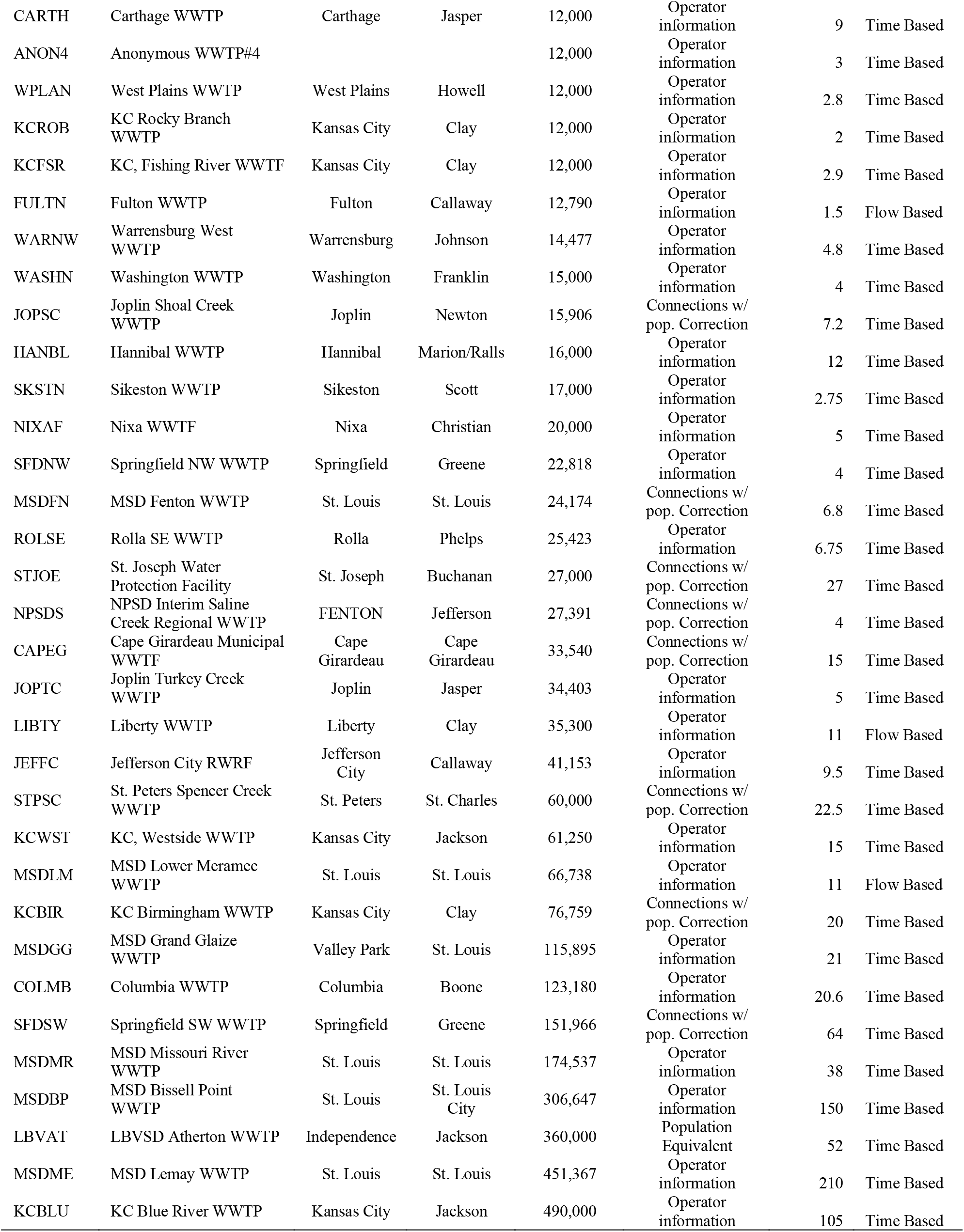
Long-time monitoring wastewater facilities across Missouri state and population sizes it served.

**Table S2.**
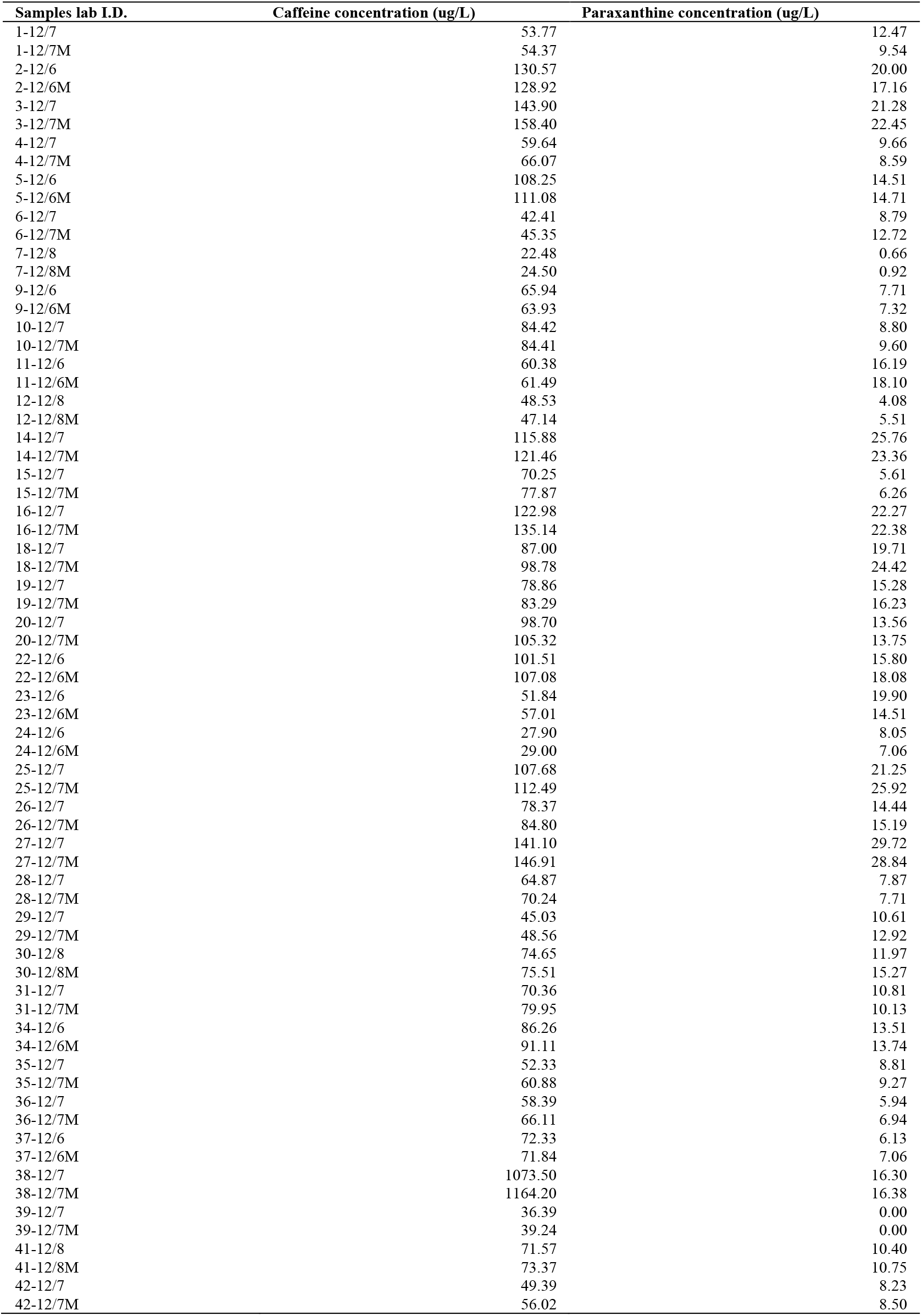
Comparison of caffeine and paraxanthine concentrations from two sample preparation methods with samples of weeks of December 6, 2021. “M” indicated dilution solution was methanol, and the corresponding sample above with the same sample I.D. and date (e.g., 1-12/7) but without “M” means method with acidification and dilution solution was LC-MSMS buffer.

| Samples lab I.D. | Caffeine concentration (ug/L) | Paraxanthine concentration (ug/L) |
| --- | --- | --- |
| 1-12/7 | 53.77 | 12.47 |
| 1-12/7M | 54.37 | 9.54 |
| 2-12/6 | 130.57 | 20.00 |
| 2-12/6M | 128.92 | 17.16 |
| 3-12/7 | 143.90 | 21.28 |
| 3-12/7M | 158.40 | 22.45 |
| 4-12/7 | 59.64 | 9.66 |
| 4-12/7M | 66.07 | 8.59 |
| 5-12/6 | 108.25 | 14.51 |
| 5-12/6M | 111.08 | 14.71 |
| 6-12/7 | 42.41 | 8.79 |
| 6-12/7M | 45.35 | 12.72 |
| 7-12/8 | 22.48 | 0.66 |
| 7-12/8M | 24.50 | 0.92 |
| 9-12/6 | 65.94 | 7.71 |
| 9-12/6M | 63.93 | 7.32 |
| 10-12/7 | 84.42 | 8.80 |
| 10-12/7M | 84.41 | 9.60 |
| 11-12/6 | 60.38 | 16.19 |
| 11-12/6M | 61.49 | 18.10 |
| 12-12/8 | 48.53 | 4.08 |
| 12-12/8M | 47.14 | 5.51 |
| 14-12/7 | 115.88 | 25.76 |
| 14-12/7M | 121.46 | 23.36 |
| 15-12/7 | 70.25 | 5.61 |
| 15-12/7M | 77.87 | 6.26 |
| 16-12/7 | 122.98 | 22.27 |
| 16-12/7M | 135.14 | 22.38 |
| 18-12/7 | 87.00 | 19.71 |
| 18-12/7M | 98.78 | 24.42 |
| 19-12/7 | 78.86 | 15.28 |
| 19-12/7M | 83.29 | 16.23 |
| 20-12/7 | 98.70 | 13.56 |
| 20-12/7M | 105.32 | 13.75 |
| 22-12/6 | 101.51 | 15.80 |
| 22-12/6M | 107.08 | 18.08 |
| 23-12/6 | 51.84 | 19.90 |
| 23-12/6M | 57.01 | 14.51 |
| 24-12/6 | 27.90 | 8.05 |
| 24-12/6M | 29.00 | 7.06 |
| 25-12/7 | 107.68 | 21.25 |
| 25-12/7M | 112.49 | 25.92 |
| 26-12/7 | 78.37 | 14.44 |
| 26-12/7M | 84.80 | 15.19 |
| 27-12/7 | 141.10 | 29.72 |
| 27-12/7M | 146.91 | 28.84 |
| 28-12/7 | 64.87 | 7.87 |
| 28-12/7M | 70.24 | 7.71 |
| 29-12/7 | 45.03 | 10.61 |
| 29-12/7M | 48.56 | 12.92 |
| 30-12/8 | 74.65 | 11.97 |
| 30-12/8M | 75.51 | 15.27 |
| 31-12/7 | 70.36 | 10.81 |
| 31-12/7M | 79.95 | 10.13 |
| 34-12/6 | 86.26 | 13.51 |
| 34-12/6M | 91.11 | 13.74 |
| 35-12/7 | 52.33 | 8.81 |
| 35-12/7M | 60.88 | 9.27 |
| 36-12/7 | 58.39 | 5.94 |
| 36-12/7M | 66.11 | 6.94 |
| 37-12/6 | 72.33 | 6.13 |
| 37-12/6M | 71.84 | 7.06 |
| 38-12/7 | 1073.50 | 16.30 |
| 38-12/7M | 1164.20 | 16.38 |
| 39-12/7 | 36.39 | 0.00 |
| 39-12/7M | 39.24 | 0.00 |
| 41-12/8 | 71.57 | 10.40 |
| 41-12/8M | 73.37 | 10.75 |
| 42-12/7 | 49.39 | 8.23 |
| 42-12/7M | 56.02 | 8.50 |

**Fig. S1.**
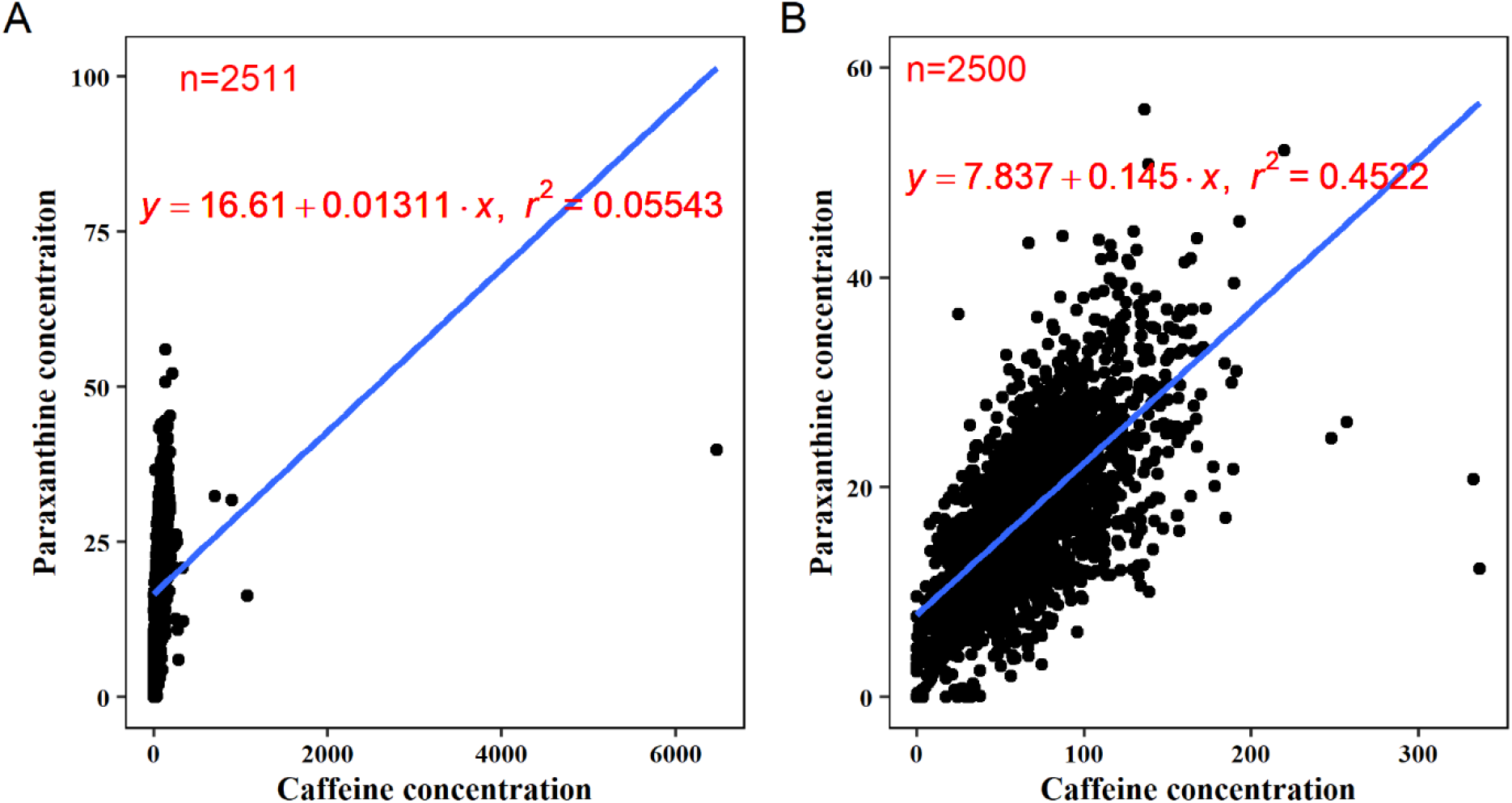
The relationship between caffeine and paraxanthine concentrations. Plot A included all the available data, but plot B was without Sikeston Wastewater Treatment Plant (SKSTN).

**Fig. S2.**
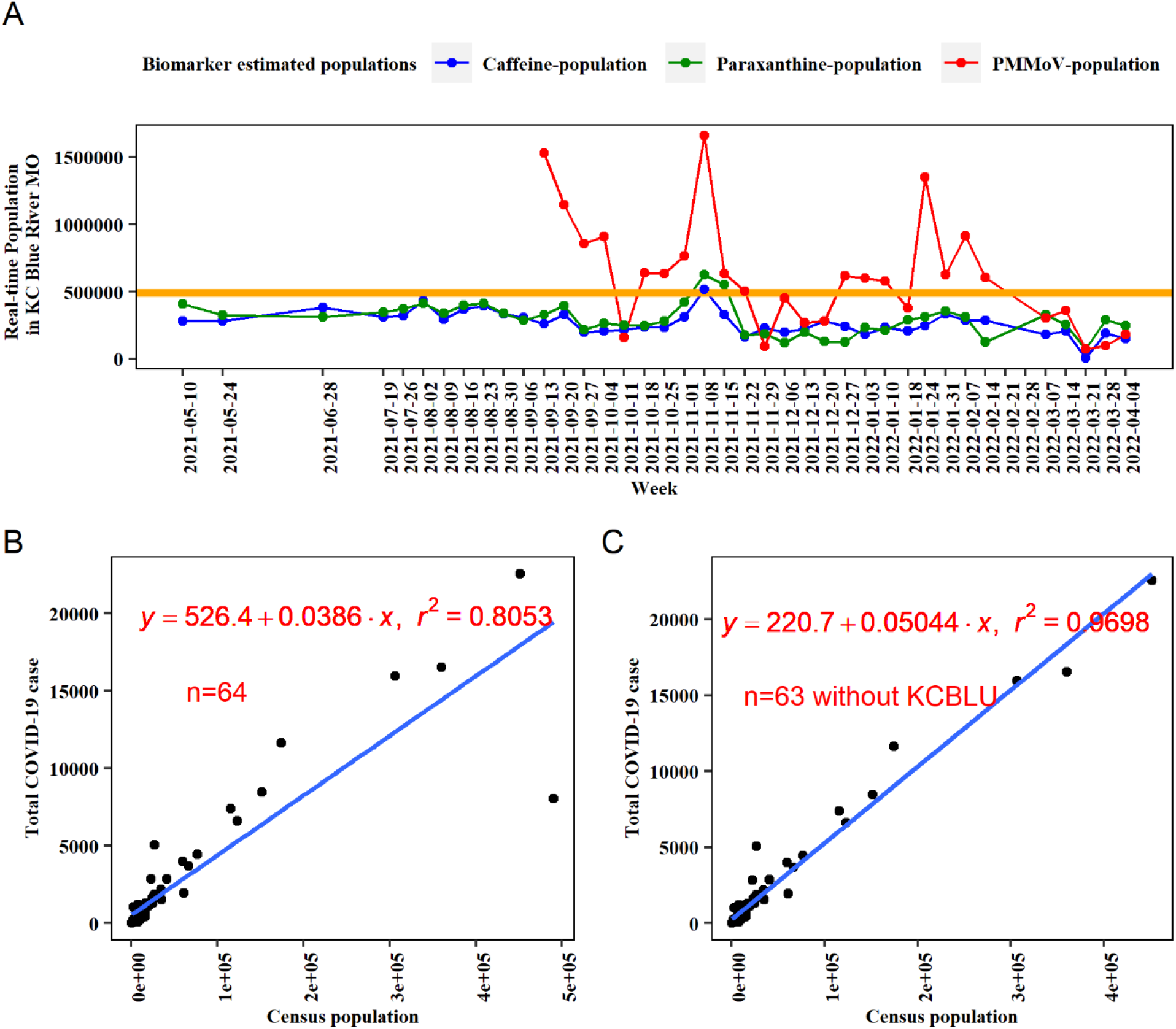
A: Predicted real-time populations in the area served by the Kansas City Blue River wastewater treatment plant (WWTP KCBLU) using concentrations of wastewater biomarkers based on the models with the data from WWTP KCBLU. The metadata population was plotted using the orange line for reference. The total Covid-19 clinical cases of each WWTP from 09/13/2021 to 04/10/2022 were strongly increased with the metadata population size (B: with WWTP KCBLU; C: without WWTP KCBLU).

**Fig. S3.**
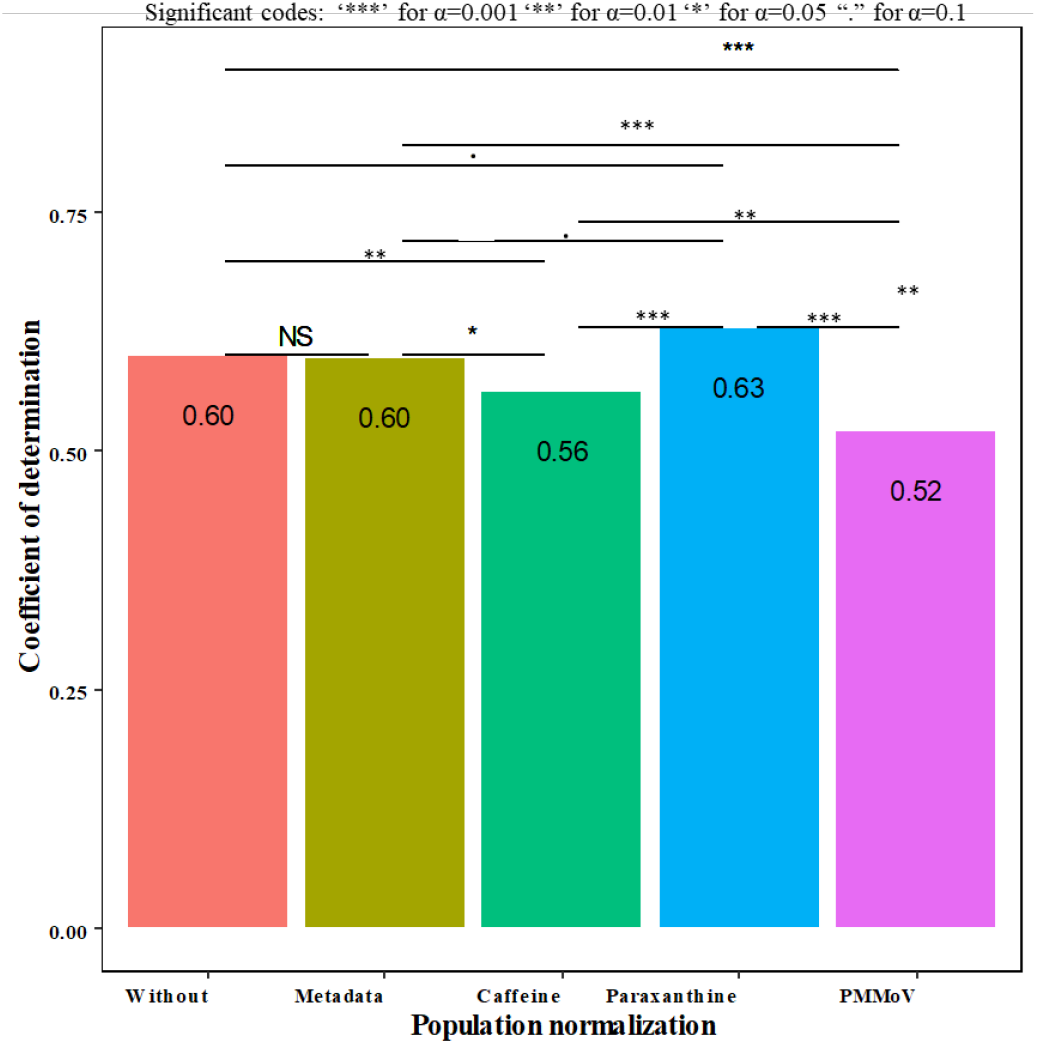
Comparison of the coefficient of determination of the linear regression models (R^2^) between wastewater SARS-CoV-2 RNA load (copies/week/10K person) and clinic COVID-19 incidence rate (cases/week/10K person) within 64 WWTPs without and with the population normalizations

**Fig. S4.**
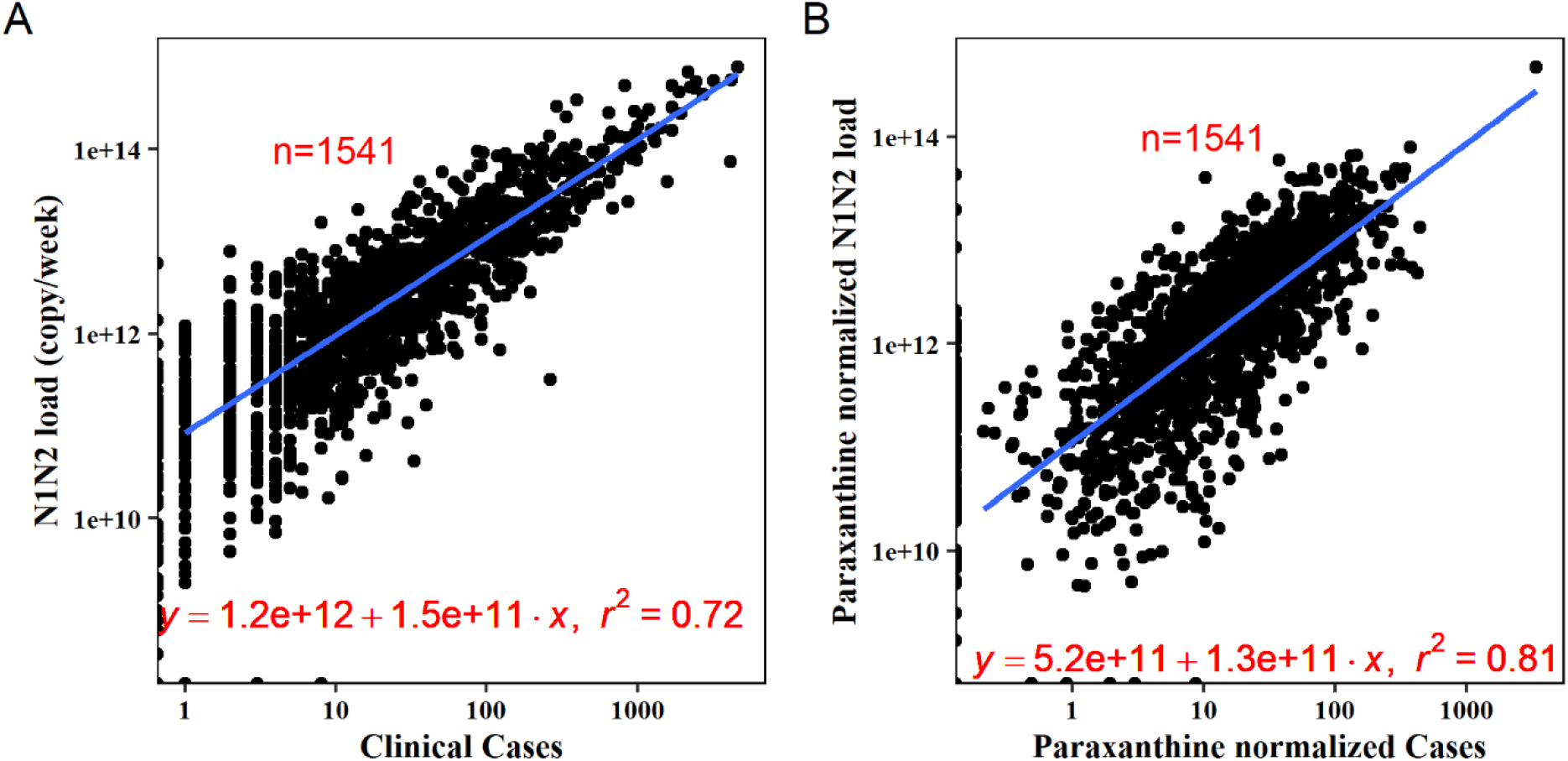
The linear regression relationship between wastewater SARS-CoV-2 RNA load with clinical cases (A: without population normalization; B: paraxanthine population normalization). The slopes of the regression equation indicated wastewater SARS-CoV-2 RNA concentration per week and person.

